# Clinical Advancement Forecasting

**DOI:** 10.1101/2024.08.02.24311422

**Authors:** Eric Czech, Rafal Wojdyla, Daniel Himmelstein, Daniel Frank, Nick Miller, Jack Milwid, Adam Kolom, Jeff Hammerbacher

## Abstract

Choosing which drug targets to pursue for a given disease is one of the most impactful decisions made in the global development of new medicines. This study examines the extent to which the outcomes of clinical trials can be predicted based on a small set of longitudinal (temporally labeled) evidence and properties of drug targets and diseases. We demonstrate a novel statistical learning framework for identifying the top 2% of target-disease pairs that are as much as 4-5x more likely to advance beyond phase 2 trials. This framework is 1.5-2x more effective than an Open Targets composite score based on the same set of evidence. It is also 2x more effective than a common measure for genetic support that has been observed previously, as well as in this study, to confer a 2x higher likelihood of success. Utilizing a subset of our biomedical evidence base, non-negative linear models resulting from this framework can produce simple weighting schemes across various types of human, animal, and cell model genomic, transcriptomic, proteomic, and clinical evidence to identify previously undeveloped target-disease pairs poised for clinical success. In this study we further explore: i) how longitudinal treatment of evidence relates to leakage and reverse causality in biomedical research and how temporalized evidence can mitigate common forms of potential biases and inflation ii) the relative impact of different types of features on our predictions; and iii) an analysis of the space of currently undeveloped, tractable targets predicted with these methods to have the highest likelihood of clinical success. To ease reproduction and deployment, no data is used outside of Open Targets and the described methods require no expert knowledge, and can support expansion of lines of evidence to further improve performance.

## 1 Introduction

It has been well established that drugs with human genetic evidence linking their respective targets to indications in clinical trials are more likely to succeed [1, 2, 3, 4, 5], typically by a factor of at least 2x [6, 7, 8, 9, 10]. This information has been used to devise target and target-disease ranking algorithms based primarily on a synthesis of multiple genetic signals alone [11, 12, 13]. It is also possible to expand the breadth of this genetic support to more targets and diseases based on knowledge graphs, protein interactions and/or disease ontologies [14, 15, 10, 16, 17]. <u>To our knowledge, all</u> <u>such expansion methods identify a larger space of opportunities at the expense of expected success rates</u>. In contrast, we aim to establish a framework for identifying target-disease (TD) pairs with the very highest possible likelihood of success in order to establish an evidence-based method to optimize research and development pipelines. This is accomplished in this study by integrating a subset of our evidence base from Open Targets [12] spanning human clinical, genomic, transcriptomic and proteomic data as well as cell and animal model evidence, pathway information, and basic literature metrics, in a simple statistical modeling framework.

This method contrasts with far more integrative methods that rely on neural and/or graph models over extensive knowledge graphs [18, 19, 20, 21], which are more complex and often difficult to interpret. We believe a desirable middle ground between these two approaches and those that aim to combine many orthogonal indicators of success through expert knowledge in heuristic systems [22, 12] would: 1) permit inclusion of many types of evidence from many sources to maximize inclusion of potentially causal, orthogonal predictive properties of targets and diseases, 2) be highly interpretable, 3) support expert judgement where necessary, and 4) not require manual ranking/weighting schemes.

<u>A substantial challenge inherent to building such a system is the need to account for the longitudinal nature of knowledge discovery in biomedicine.</u> This is vital because any method that optimizes for likely *future* clinical success based on *historical* clinical outcomes may easily be biased by the non-random nature with which evidence can be absent. By utilizing “temporalized” evidence – i.e. evidence for which timing of its emergence can be determined – and outcomes when training our methods before ultimately applying them to present-day evidence with no restrictions on timing, we reduce the risk of information leaks and inflation of performance estimates. We discuss motivations, prior research and our own analysis on how important this problem is for each source in Section 2.7.

Solutions to such problems are common, but not fully addressed even in precedent studies by other groups that attempt to predict clinical trial outcomes using similarly temporalized predictors [23, 24, 25]. The need for longitudinal features is most clear in settings where the inclusion of predictors like historical success rates for targets or diseases, trial sponsor track records, eventual patient enrollment, or other time-based events and outcomes, would otherwise constitute information leaks. This is discussed in some depth in [23] which notes several studies that do not account for this problem, and presents both quasi-prospective as well as prospective results. The difference between the two is that the former reconstructs timelines for predictors and outcomes based on recorded event dates while the latter relies on frozen predictions that are never evaluated until years later. Nomenclature for these formulations is conflicting though, where this definition of a quasi-prospective design is deemed entirely prospective in some cases (e.g. [26]). To avoid any potential confusion, our formulation in this study is quasi-prospective. The distinct advantages and disadvantages of each design provide a complementary risk/reward trade-off, and we assert that the distinction between them is important. This is discussed more in Section 4.

Precedent studies and methods discussed so far can largely be categorized as either 1) target and target-disease prioritization methods evaluated based on how well they correlate with observed clinical trial success, and 2) clinical trial outcome prediction models. Both are measured against the same outcomes and an important distinction between them lies in how prioritization methods are **not** directly optimized for those outcomes while outcome prediction methods are. <u>In this study, we attempt to bridge these methodologies by predicting clinical trial advancement for target-disease</u> <u>pairs based solely on information that would be present well in advance of any drug program or individual trial</u>. We then calibrate these predictions to determine what thresholds are necessary to match the observed success rates from benchmarks for genetic support like OMIM [27], ClinVar [28] and GWAS. Finally, we examine how many present-day target-disease pairs are likely to see success rates matching or exceeding those calibrated benchmarks, that are also undeveloped (i.e. have never been in clinical trials) and have a tractable target (i.e. with evidence suggesting that therapeutic modulation is possible).

## 2 Results

In order to model the probability of clinical success, or “clinical advancement” for target-disease (TD) pairs, we first define “advancement” as progression beyond any particular trial phase across all drugs associated with a single TD pair. We consider an advancement to have occurred when a later-stage trial is observed for the same pair. When no later-stage trial is observed, no advancement is assumed and this binary condition provides the outcome to predicted using the features shown in Table 1. Due to the lack of timestamped drug approvals in Open Targets, all study results presented here consider only advancement beyond phase 2. This limitation is discussed further in Section 6.

Information for each of the features in Table 1 that are denoted as temporal is only used when it was published in a year **prior** to the first phase 2 trial for a TD pair. Genetic evidence is a notable exception to this rule, and motivations for that exception are detailed in Section 2.7. A training dataset is then formed by including only TD pairs where the first phase 2 year is between 1990 and 2015. The evaluation dataset then consists of all TD pairs entering phase 2 between 2016 and 2022, with a 2 year offset from the present year (2024) to allow enough time for some trials to complete. While the average phase 2 trial duration may be as low as 2 years [29], other estimates would suggest half of them take longer than 2.9 years [30]. This means a substantial fraction of recent outcomes are censored, that this is an important parameter to test sensitivity to and that time itself is likely to be a crucial covariate in this formulation. The distribution of these outcomes, the number of associated targets/diseases and a variety of other statistics on this dataset are presented in Supplementary Figure 8.

### 2.1 Features

The features used throughout this study consist of 27 target-disease pair predictors, 5 target-specific predictors and 1 disease-specific predictor – a subset of our full productionized feature set. These are listed in Table 1. The target and disease specific features are chosen carefully such that they are either capable of being associated with years in which events supporting them occurred or result from large-scale, unbiased methods. This choice was motivated in part by empirical results presented in Section 2.7 and assumptions described more in Section 5. Simply put, the data in this study combines scores from Open Targets for target-disease evidence and a select subset of target prioritisation [32] fields with almost no modifications, other than to add target and disease specific indicators of maximum trial phases reached and two extra genetic association features (also described in Section 5).

The combination of target, disease and target-disease features creates sparsity patterns that are important to understand before interpreting models built from it. Figure 1 demonstrates this sparsity by showing that of all the TD pairs entering phase 2 trials for the first time between 2016 and 2022 in our evaluation dataset (N=9010), slightly less than 2% of them (1.89%) ever have evidence directly linking them other than literature co-mentions, which exist for 21% of those pairs. These TD pairs do, however, very frequently have prior clinical evidence for their associated targets and diseases. Specifically, 8,425 (94%) have a target and 6,855 (76%) have a disease that had already been in phase 2 or later trials previously. This is consistent with herding effects observed in recent drug development pipelines [33] over the same time period (2016-2022) and underscores the prognostic value such information may have as it is becoming more and more common and clearly confers lower clinical risk for new drug programs. We also observe that, on top of the clear theoretical, causal relationship between prior target and disease human clinical validation and the likely success of programs for new combinations of such targets and diseases, these features have strong, univariate predictive effects in the evaluation dataset of our study. This is illustrated in Supplementary Figure 10, which shows the relative risk of advancement for these features capturing the highest clinical stage previously reached by a target or disease.

**Figure 1:**
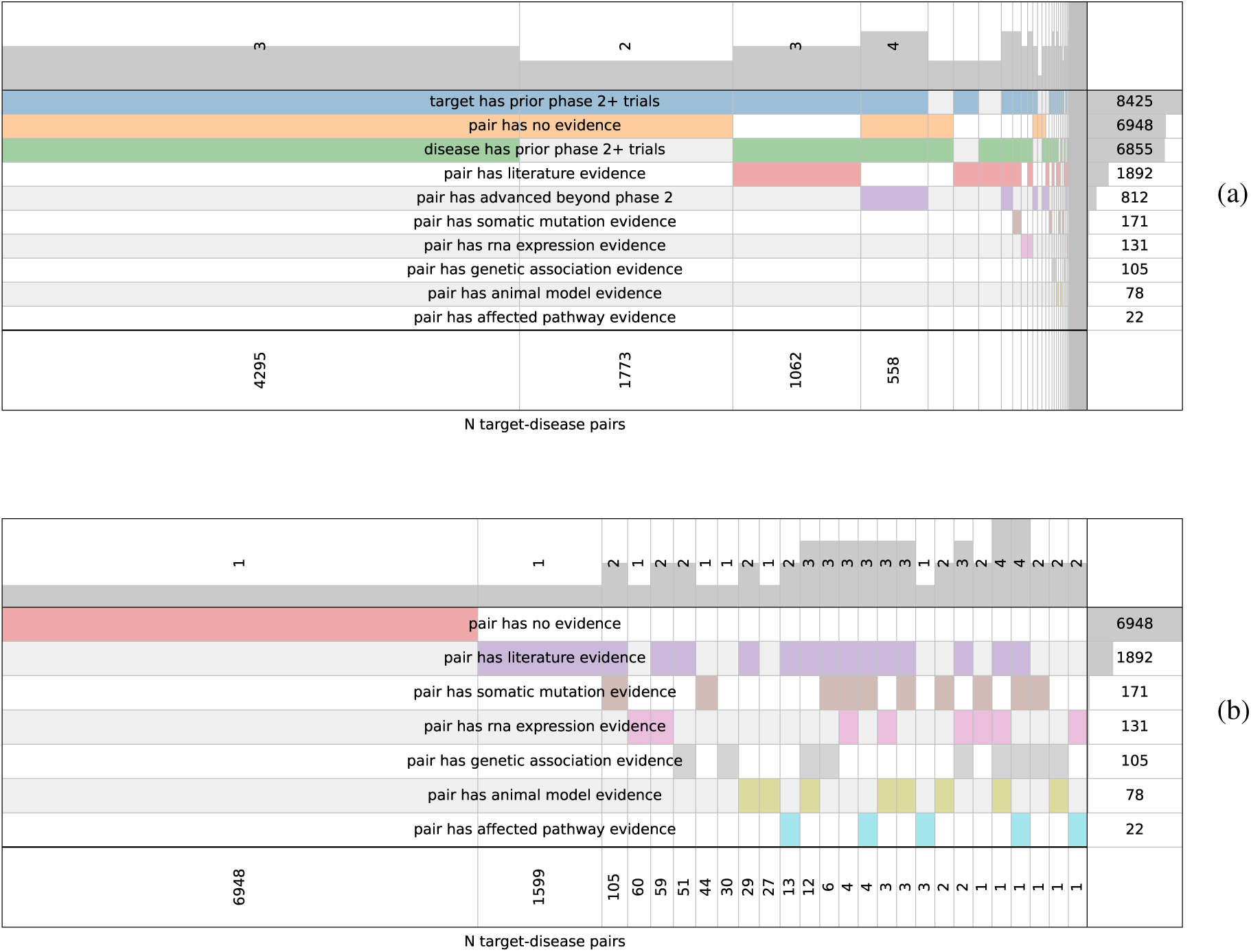
Evaluation dataset feature presence. (a) Presence and co-occurrence of select features for overlapping sets of TD pairs. Each column of the visualization indicates a distinct grouping of TD pairs for which one combination of features is present. The rows denote features and the existence of any color bar other than white or grey indicates that the feature is present. For example, the first column from left-to-right shows that there are 4,295 TD pairs in the evaluation dataset with **only** target-specific and disease-specific clinical evidence (phase 2+ trials); i.e. no evidence exists that is specific to the TD pair for those cases. The size of each bar within a column is proportional to the number of TD pairs with that combination of features, and the sort ordering is determined by the number of TD pairs associated with any one feature (show in the right margin). The top margin shows the number of features in any one feature combination and the bottom margin shows the number of TD pairs that are a part of that same combination. There are 9010 TD pairs in the evaluation dataset overall. (b) Same as (a) with proportional bar scaling that does not drop beyond a limit necessary to fit all margin labels and combinatorial groupings. Features that are not specific to TD pairs are omitted for brevity, along with the outcome feature used in this study and denoted here with the label “pair has advanced beyond phase 2”. For more details on the origin and interpretation of these plots, see supervenn [31].

Taken together, the paucity of TD pair evidence and the abundance of prior target or disease clinical validation should be considered carefully when interpreting predictive performance. TD pairs predicted to be highly likely to advance clinically despite a lack of target-disease-specific evidence are entirely plausible, but the utility of such a prediction may vary depending upon the application. We choose to minimize the influence of these cases in our study by focusing on rankings within therapeutic areas that do not extend beyond the number of TD pairs with direct evidence of some kind, as discussed in Section 5. This choice is also reflected in our primary performance metrics, as discussed in Section 2.3.

### 2.2 Models

We develop our predictive framework using two primary types of models: linear models, where relationships between features and outcomes are assumed to be linear, and gradient-boosted tree models, which can capture non-linear relationships. Both models were tested in constrained formats, ensuring all feature effects are non-negative, and unconstrained formats, allowing for negative relationships. This is possible with no underlying feature transformations because all scores in Open Targets are constructed such that higher scores are presumed to be advantageous. We also note that neural models are a significant omission from this framework. While deep learning methods for tabular data are competitive with gradient-boosted tree models [34, 35, 36], they are rarely superior without transfer learning [34, 37, 38]. Additionally, the data in this proof-of-concept is limited to Open Targets evidence scores that already reflect a large degree of feature engineering, thereby limiting the extent to which deep learning networks can identify more granular predictive signals.

In order to assess the value of groups of related features, we also apply these models to our evaluation dataset using several feature ablations. We refer to a “core” feature set consisting of all features listed in Table 1 except for the sole feature capturing the time since a target-disease pair first entered phase 2 trials (i.e. target_disease__time__transition). Combinations of learning algorithms for the models and the feature sets to which they are applied are referred to using the following convention:

- **RDG**: Constrained, L2-regularized linear regressor (a.k.a. “Ridge regressor”) fit with all core features
- **RDG-T**: **RDG** fit with all features instead of only core features, where the only difference is the inclusion of time since phase 2 transition for a TD pair
- **RDG-X**: **RDG** fit *without* human clinical and genetic evidence features
- **GBM**: Constrained, gradient-boosted machine with fit with all core features
- **GBM-T**: **GBM** fit with all features instead of only core features

The omitted human clinical and genetic evidence features for the RDG-X model are all of those in Table 1 with the midfix “clinical” or “genetic_association”. We omit these features specifically because they are known or expected to comprise good predictors of human clinical success, so their exclusion examines the extent to which only literature and animal model evidence along with target-specific properties accomplish this task.

In order to compare these models to an Open Targets composite score (OTS), we use an equally weighted sum of all scores from individual sources except for those assigned lower weights in [39]. Scores from these sources are multiplied by the corresponding weight before being summed and only the target-disease-specific features of Table 1 are used. Neither the target-specific and disease-specific features nor the time since phase 2 transition feature are included in this calculation.

### 2.3 Metrics

The primary performance metric used in this study is relative risk (RR). This metric is commonly used to assess univariate measures of genetic support [7, 8, 6] and can be more intuitively understood, in the context of this study, as the probability that a TD pair among the top *N* TD pairs as ranked by a particular method will advance beyond phase 2 trials divided by that same probability of advancement among TD pairs with a rank greater than *N*. This provides a means to compare multivariate, model-based methods to univariate methods on a common scale. More specifically, any RR metric reported for a model among top *N* rankings is defined as:

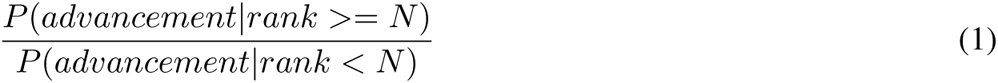

and RR metrics reported for univariate methods based on Open Targets scores for a single type of evidence, where not stated otherwise, are defined as:

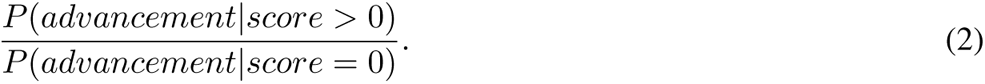

The use of such a metric is essential for properly assessing performance in this forecasting problem. While we also report more common measures of classifier performance like Receiver Operating Characteristic (ROC) and Average Precision (PR), neither of these adequately capture behavior in the upper extremes of rankings due to the sparsity with which TD pair evidence is present for pairs that have ever entered phase 2 trials. This sparsity is further exacerbated in this study by the constraint that most of that evidence must have existed *before* such trials began. For more details on the extent of this sparsity, see Section 2.1. The figure presented there, Figure 1, also demonstrates that a substantial fraction (78%) of TD pairs that ultimately advance beyond phase 2 trials have targets and/or diseases with prior clinical validation despite no direct evidence linking the pairs themselves, which means a comprehensive measure of classifier performance (e.g. ROC) is far more likely to reflect the extent to which disease-only historical, clinical information or target-only information – including other attributes like conservation and tissue expression – can predict clinical success.

Again, this is not our primary focus as we want to evaluate the maximum achievable performance in this forecasting problem, and we assert that this is best accomplished when one or more lines of target-disease-specific evidence are present.

Reasonable alternative choices for this primary metric include those more common in information extraction literature or other machine learning studies with a focus on ranking rather than classification, such as mean reciprocal rank (MRR), precision at *k* (P@k) and normalized discounted cumulative gain (NDCG) [40, 41]. Precision at *k* is the most similar among these to relative risk at *k* since it is equivalent to the numerator in the relative risk calculation. We use relative risk instead because it is more intuitive than most ranking metrics, has well established analytical solutions for confidence intervals [42] and is consistent with prior work in this field.

Lastly, we emphasize that the interpretation of “risk” for the relative risk metric is to be inverted in this context. A higher “risk” in this study actually corresponds to a greater probability of success. The name “Relative Success” is used for this metric instead in [6] even though it has the same underlying definition. We choose not to use this label because we also present generic performance measures like ROC and AP, thereby prioritizing consistency with a domain-independent nomenclature.

### 2.4 Performance

#### 2.4.1 Open Targets comparison

Figure 2 demonstrates how well our primary model in this study, RDG, ranks TD pairs by comparison to a composite score from Open Targets, OTS. This comparison highlights relative risk (RR) as our primary performance indicator along with secondary measures of performance like Receiver Operating Characteristic (ROC) and Average Precision (PR), as discussed more in Section 2.3. We observe improved performance over this benchmark across all metrics, and provide further comparisons between the OTS score and other models in Section 2.4.3.

**Figure 2:**
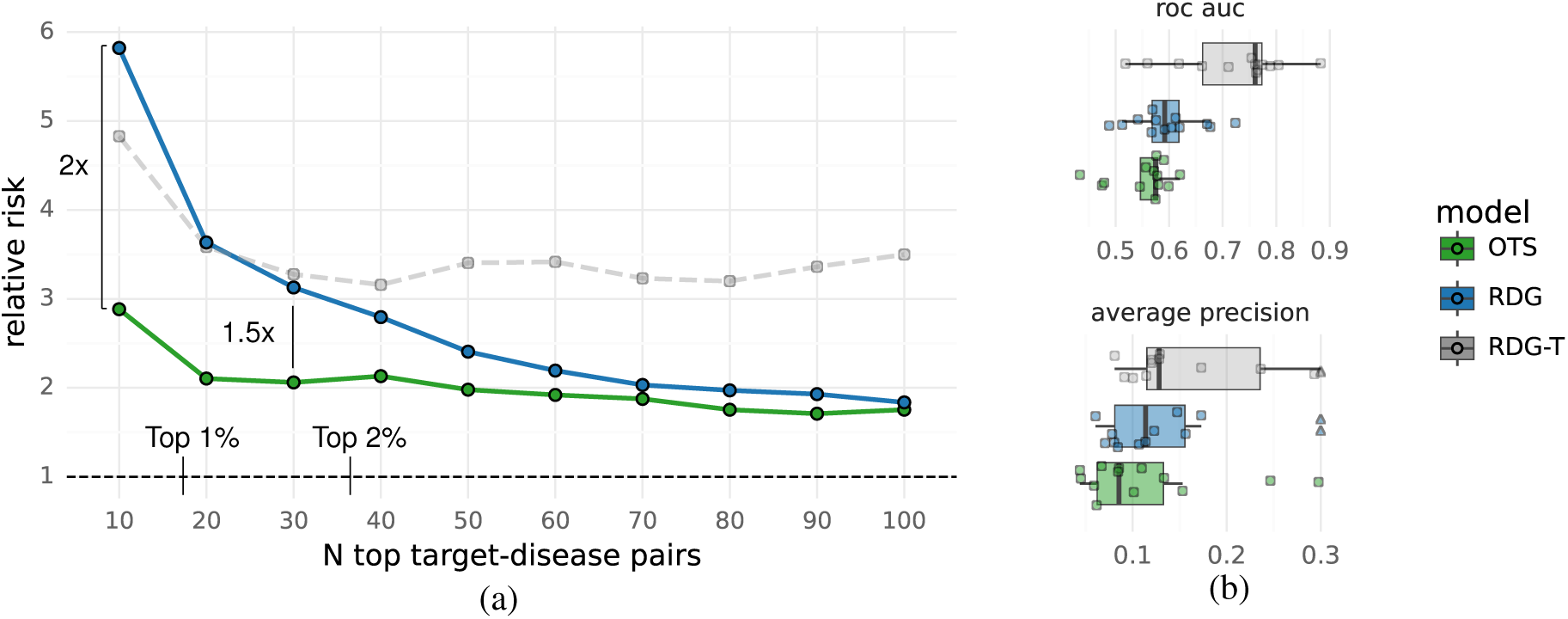
Performance compared to Open Targets composite scores. (a) Equally-weighted average relative risk estimates across 13 therapeutic areas, by number of top rankings and 3 methods: RDG (ours), RDG-T (ours) and OTS (Open Targets composite scores). (b) Receiver operating characteristic (ROC) and average precision scores across the same 13 therapeutic areas (each dot is one therapeutic area) with no limit on the number of rankings. See Supplementary Figure 12 for raw data underlying (a). Annotations indicate RR multiples between RDG and OTS for the top 10 and top 30 TD pairs as well as how many TD pairs comprise the top 1% (N=18) and top 2% (N=36) of rankings on average across therapeutic areas. All results shown are from the evaluation dataset.

The third ranking method presented in Figure 2, “RDG-T”, differs from the RDG model only in that it uses time since the phase 2 transition as a predictive factor in addition to all others. We observe that the use of this information greatly improves standard performance metrics like receiver operating characteristic (ROC) and average precision (AP), however it adds little to no value in rankings beyond a level where substantial relative risk increases can be observed. In other words, it constitutes an effective but coarse mechanism for ranking TD pairs while lacking the high precision of other factors like genetic support. More implications of this and opportunities it may imply are discussed in Section 4. As a more practical concern, we refrain from focusing on RDG-T, or the similar GBM-T model, because neither is readily applicable to undeveloped TD pairs for which the time since phase 2 transition is not available. They do, however, present a useful performance ceiling towards which future work might build.

We also note that Figure 2 presents average RR estimates drawn across a subset of therapeutic areas, and the criteria used to select them is described more in Section 5. A full list of therapeutic areas meeting these criteria can be seen in Supplementary Figure 12 along with the RR estimates summarized in Figure 2. Furthermore, a comparison of the distribution of these estimates by model is presented in Supplementary Figure 11 along with the statistical significance of their differences.

We conclude that the RDG model outperforms OTS by a factor of at most 2 and least 1.5 for a significant number of TD pairs. A sensitivity analysis for this key result is presented in Supplementary Section 9.1, where we find that these multipliers hold on average across multiple Open Targets releases and other important settings.

#### 2.4.2 Genetic benchmark comparison

In order to establish baseline levels of success and coverage across TD pairs, we examine ranking performance in comparison to well established, univariate indicators of genetic support in Figure 3. This figure presents OMIM and GWAS baselines, in the parlance of [8], [7] and [6], as well as an intermediate baseline from the European Variation Archive (EVA) [43] containing evidence predominantly from ClinVar [28].

**Figure 3:**
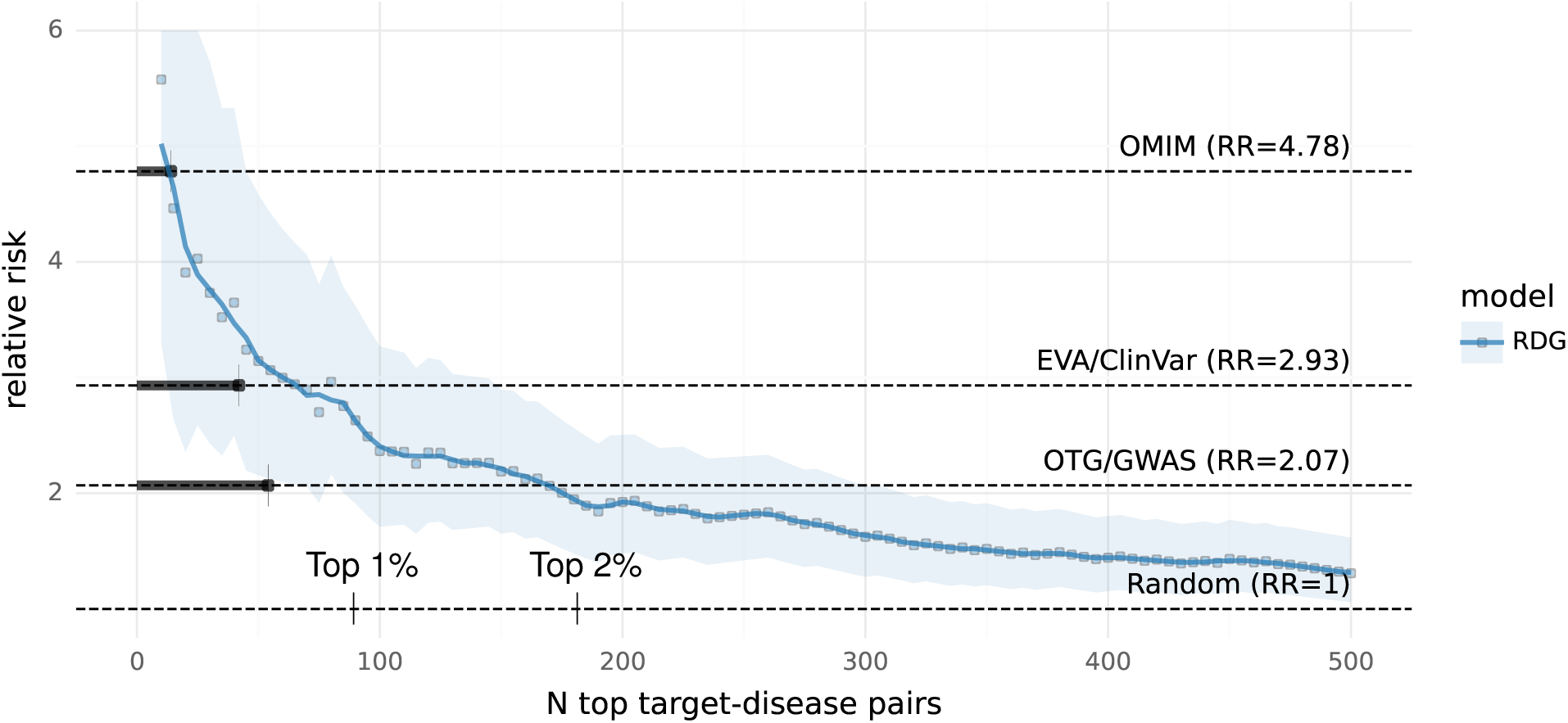
Performance compared to genetic support benchmarks. The RR estimates for each benchmark annotated on the right are shown as horizontal dotted lines and are calculated across all TD pairs (N=9010). The number of pairs with support from each benchmark is represented by the bars extending horizontally (left to right) from the y-axis. RDG RR point estimates are shown in blue and bounds around the estimates correspond to Katz 90% confidence intervals. Annotations on the x-axis indicate how many TD pairs comprise the top 1% (N=90) and top 2% (N=180) of rankings. All results shown are from the evaluation dataset.

One key objective of this study is to determine if any model, e.g. RDG, can sort TD pairs with genetic support such that at least some portion of that sorted list has a likelihood of advancement that consistently exceeds what is expected from any one source of genetic support. We find that this goal is met and surpassed by the RDG model, which actually identifies more TD pairs than those that have either EVA or GWAS support and have an equivalent or greater expected rate of advancement. See Section 2.6 for more on how these benchmarks are employed to contextualize opportunities among undeveloped TD pairs and Supplementary Figure 13 for top predictions from the RDG model. This latter, supplementary figure further emphasizes our focus on prioritizing opportunities beyond those with genetic support and provides examples of TD pairs with multiple lines of evidence.

We also note that Supplementary Figure 9 shows confidence intervals for each of the genetic benchmarks of Figure 3 in isolation, as well as all other target-disease-specific evidence sources, in addition to confidence intervals for the RDG model at various top ranking cutoffs. Similar comparisons for target-specific and disease-specific features can be seen in Supplementary Figure 10.

#### 2.4.3 Model comparison

Figure 4 presents average performance across therapeutic areas for select combinations of learning algorithm, constraint type and feature group described in Section 2.2. Several key findings illustrated in this figure are:

1. The RDG-T model achieves far higher ROC and AP scores through the use of the time since transition feature, which indicates the number of years a TD pair has been under development after having reached phase 2.
2. The RDG model, however, matches or exceeds RDG-T in performance among top TD pairs
3. The RDG-X model, using no human clinical or genetic evidence linked to a disease, outperforms the Open Targets composite score and nearly matches the performance of the RDG model beyond top rankings
4. Linear models outperform gradient-boosting models by nearly all measures
5. Constrained linear models outperform unconstrained linear models by nearly all measures

**Figure 4:**
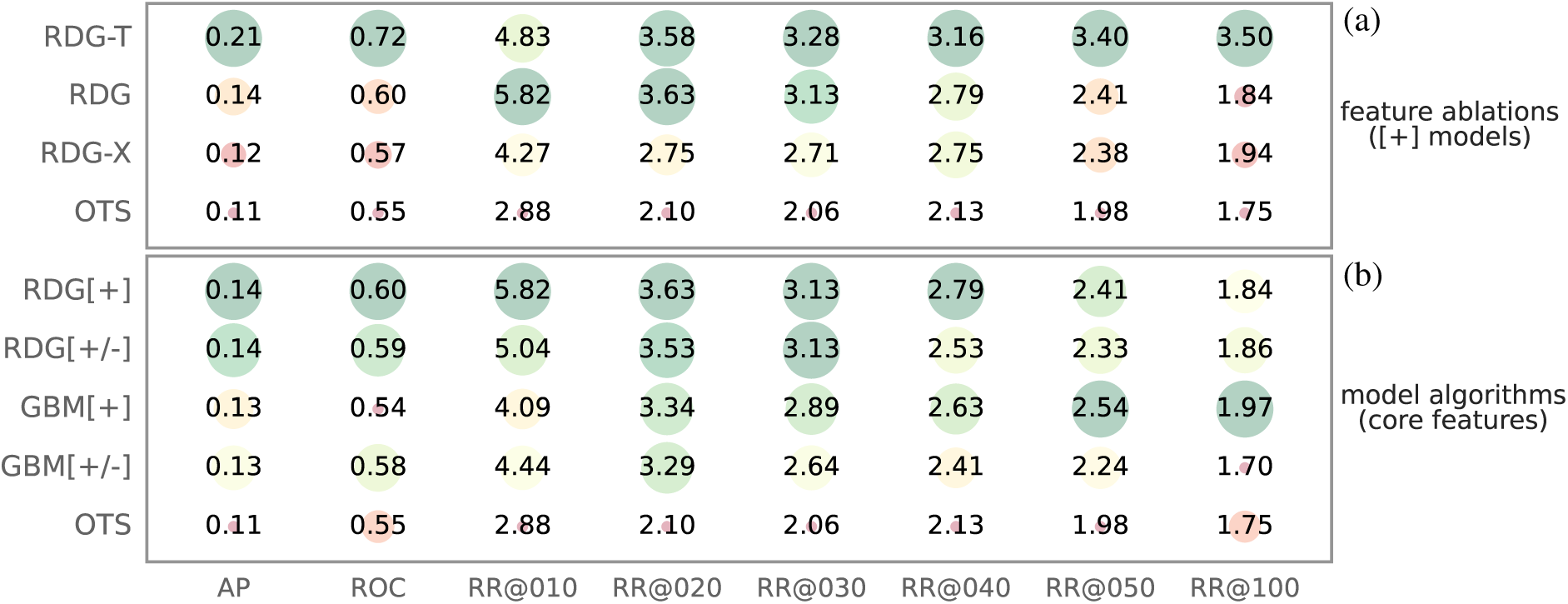
Performance across model algorithms and feature ablation groups. Average precision (AP) and receiver operating characteristic (ROC) scores with relative risk (RR) at ranking cutoffs denoted by RR@N. Dot sizes and colors (red=low score, green=high score) are both set according to the score shown in each cell. (a) Performance across constrained RDG models using different groups of features as described in Section 2.2 (b) Performance across constrained ([+]) and unconstrained ([+/-]) linear and gradient-boosted models using the core feature set.

We conclude from these results that constrained linear models are an optimal choice for this problem due both to their greater performance and interpretability. This interpretability is illustrated more in Section 2.5 and owed much to the effort Open Targets has already undertaken to construct evidence scores such that they can be assumed to have a monotonically increasing effect on the likelihood that a causal relationship exists between a target and a disease.

### 2.5 Feature effects

The coefficients learned by the RDG model, and the average effects they have across the evaluation dataset, are shown in Figure 5. This model most highly prioritizes genetic signals that have the greatest coverage, i.e. associations from GWAS studies through the ot_genetics_portal feature and associations from any curated clinical genetics source, i.e. EVA, Orphanet, UniProt, Genomics England, ClinGen and gene2phenotype, via the curated feature. Notably, literature and target/disease specific clinical features also have substantial effects, followed by indicators of animal evidence and target genetic constraint or expression specificity. Any features not shown were deflated to have no effect, which is possible in this model due to the non-negativity constraint. One such feature worth emphasizing is transcriptomic evidence from Expression Atlas. We found this somewhat surprising, but it is supported by arguments against transcript over/under expression as an indicator of genes that influence disease rather than the other way around [44].

**Figure 5:**
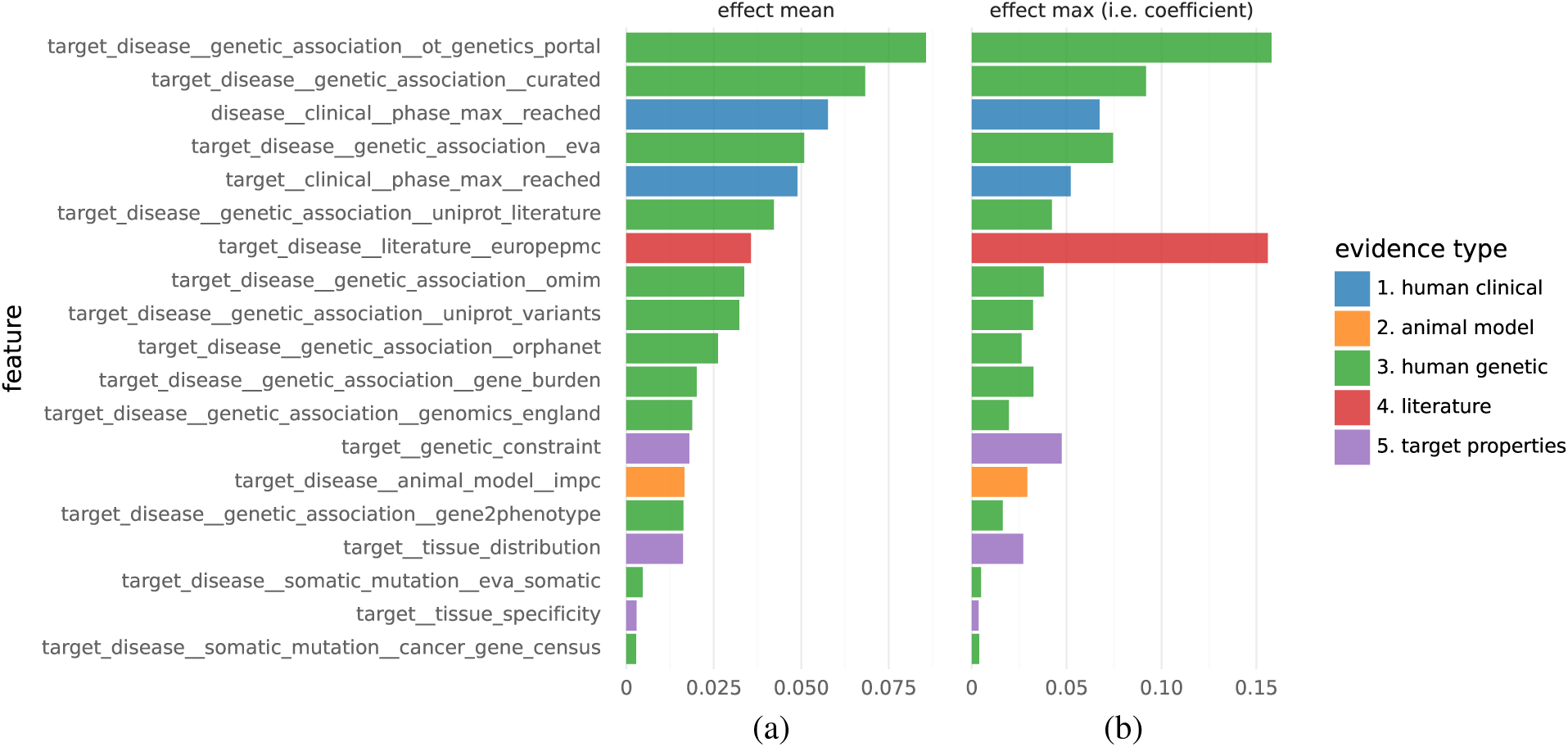
RDG model feature effects. (a) Feature average effects calculated as the mean of the product between a coefficient and a particular feature value, when that feature is present and non-negative (i.e. when it has any influence). Only the evaluation dataset is used to compute this average. The prefixes target, disease and target_disease are used to denote features that are specific to targets, diseases and target-disease pairs (respectively). (b) Feature coefficients, which are equivalent to the maximum effect that a feature can have due to all features being defined on the range [0, 1] or rescaled to it.

It is worth noting that the discordance between the coefficients and the average feature effects of Figure 5 arises from both the frequency with which features exist and the distribution of their underlying scores. Scores for many clinical genetics features (e.g. OMIM, Genomics England, UniProt) are very frequently absent or close to 1. By comparison, scores for literature associations are typically far lower, even when limited only to cases where they exist, with a median value of 0.12 (mean=.23) in the evaluation data. This is why the europepmc feature has a relatively large associated coefficient, but a much smaller average effect across predictions.

### 2.6 Rankings

Utilizing our predictive models, we systematically identify target-disease pairs that have not yet been explored in clinical trials but exhibit a high likelihood of successful outcomes. This process involves ranking pairs based on their predicted success rates and filtering those with substantial supporting evidence but no prior clinical development attempts.

A common method for identifying druggable opportunities within a specific disease context involves first ranking TD pairs according to some prioritization methodology followed by filtering or re-prioritizing those ranks based on knowledge of target tractability [45, 46, 13]. We use a similar approach to identify tractable targets associated with TD pairs that have yet to enter clinical trials. To aid in interpreting this approach, we also draw on the results of Figure 3. The data in this figure suggests thresholds for the RDG model that align to expected rates of advancement compared to several genetic support benchmarks. These thresholds are used to bucket undeveloped TD pairs before further bucketing them based on levels of tractability. The tractability buckets in Supplementary Table 2 provide HIGH, MED, and LOW confidence ratings for each type of tractability evidence based on the priorities suggested in [47].

Figure 6 demonstrates this process visually, starting from the space of all present-day TD pairs. This funnel implies there are thousands of TD pairs that 1) have never been tested in clinical trials before, 2) have a 3x (RR=2.93) higher likelihood of advancing past phase 2 trials and 3) have a target that **has** been tested clinically (in at least phase 1 trials) for one or more of the modalities highlighted. In other words, there are *∼*2,400 small-molecule-enabled, *∼*1,400 antibody-enabled, and 14 PROTAC-enabled TD pairs at the bottom of the funnel where the implied opportunity is greatest. Examples of antibody-enabled TD pairs are shown in Supplementary Figure 15 along with their corresponding genetic and clinical support. Note that these are intentionally not curated or filtered in order to capture rankings as they would be seen for the corresponding version of Open Targets (23.12). This is discussed more in Section 6.

**Figure 6:**
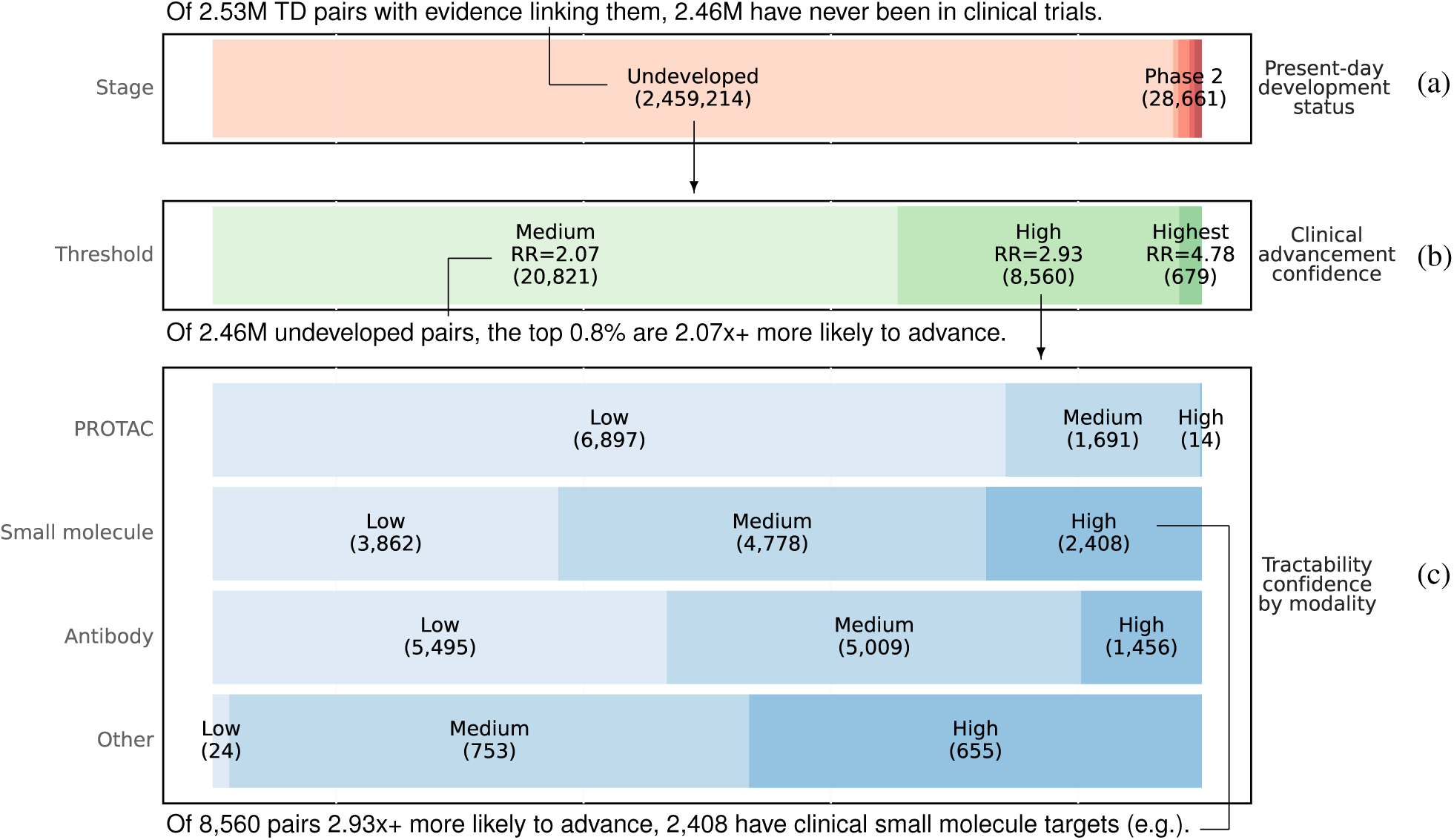
Present-day target-disease pair ranking funnel. (a) All 2.53M TD pairs in Open Targets with evidence of some kind grouped by the maximum clinical trial phase reached for each as of the present day. The Undeveloped pairs have never been tested in clinical trials. TD pair counts are shown in parentheses. Labels for phase 1, 3 and 4 trials are omitted for brevity. (b) All 2.46M undeveloped TD pairs for which RDG model predictions exceed the thresholds determined by the corresponding genetic benchmarks in Figure 3. The Medium threshold was calibrated by genetic support across OTG (i.e. GWAS evidence). The associated RR of 2.07 implies that pairs with RDG predictions above this threshold are at least 2.07 more likely to advance clinically than those below the threshold. The same is true for the High and Highest thresholds, which correspond to thresholds determined by EVA and OMIM, respectively. These groupings are not mutually exclusive, e.g. the 20,821 TD pairs above the Medium threshold are a superset of the pairs above the High threshold. (c) All 8,560 undeveloped TD pairs with an expected clinical advancement rate of at least 2.93x grouped by tractability modality and confidence level. The Other modality corresponds to any modality not shown that is associated with a target tested in clinical trials previously. The High confidence grouping for this category means those targets have approved drugs. For each other modality, the High confidence level corresponds to **any** level of clinical validation for drugs of that modality, not just approval. See Supplementary Table 2 for the definition of all modality confidence levels.

For similar statistics by therapeutic area, rather than across all of them as in Figure 6, see Supplementary Figure 14. This also shows TD pair counts for trial phases omitted from Figure 6.

Lastly, we note that these TD pairs are identified using predictors that are not constrained by temporalization. In other words, the predictions used to identify **present-day** TD pairs with a high likelihood of clinical success are based on more evidence than what is available in other sections of this study. This is primarily because some evidence in Open Targets is not associated with publications, which prohibits us from associating it with a timestamp. Avoiding that limitation is appropriate in this context.

### 2.7 Inflation

To address potential biases and inflation in our dataset, our methodology specifically considers the temporal emergence of evidence. By doing so, we ensure our predictions are grounded in data reflective of genuine biomedical insights, rather than artifacts of research focus or historical data accumulation trends.

Like most studies of this kind, we assume a “closed-world” [18] over the space of target-disease pairs and any evidence between them. This means that we do not differentiate between evidence that an association for any one pair truly does **not** exist (or is too weak to be relevant under the omnigenic model [48]), and the lack of any attempt to find that evidence in the first place. This also means that our estimate of the prognostic value for any one evidence source is subject to historical trends in biomedical research and the myriad ways that this research can be biased towards particular targets and diseases. We avoid attempting to comprehensively survey these biases in favor of offering an illustrative list of specific examples that are relevant in this study:

1. Mendelian randomization research is biased towards cardiovascular diseases as they have a disproportionate number of known, modifiable exposures [49]
2. Putative protein interactions that do not result from genome-scale or otherwise unbiased assays result in an overrepresentation of successful drug targets in resources like STRING [50], thereby inflating the success of network expansion methods over these databases to identify such targets [10].
3. Transcript expression studies run in late-stage clinical trials for a single indication, e.g. [51] linking SLE to IFN genes, are a degenerate indicator of advancement beyond earlier stage trials when the timing of this evidence is not accounted for.
4. Targets tested against more indications in clinical trials enrich for failures because the marginal cost of testing more indications decreases, but the evidence for these indications is often weaker [14].
5. Herding effects in pharma R&D pipelines around particular drug targets are becoming increasingly clear over time [33] and generate an excess of clinical evidence for those targets.

We also note that the skew in basic drug target research towards those that already have rich annotations and well characterized molecular function [52], the disproportionate representation of particular target families in pharma R&D pipelines [53, 54] and the fact that literature is well known to be biased away from negative results in general [55] are all problematic.

While it is not possible to address all of these issues, we emphasize that there is a clear pattern across the examples in the list above in that they require **past** clinical successes and/or failures to arise in the first place. <u>This suggests that accounting for when evidence first emerged would limit the extent of these problems</u>. We do so in this study based solely on publication dates associated with any one piece of information linking target-disease pairs. <u>This also offers a novel opportunity to attempt to quantify what kind of evidence suffers most from these biases</u>. Figure 7 presents results for this based on a relative risk statistic defined as:

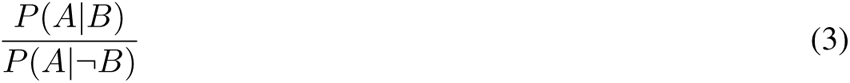

where:

- *A* is the event that evidence for a TD pair arises after its first early-stage (phase 1 or 2) trial rather than before
- *B* is the event that a TD pair advances into late-stage trials (phase 3 or 4)

We refer to this as “inflation risk” so as not to confuse it with the relative risk statistic used in all other contexts, and it can be more simply described as the fraction of TD pairs for which evidence arises **after** the beginning of an ultimately successful early-stage trial divided by that same fraction for TD pairs that do not advance to late-stage trials. The intuition for this statistic is that it will be higher if successful trials lead to the generation of evidence of a particular type, and it should be 1 in cases where the emergence of evidence is independent of clinical success. We also measure this potential lack of independence through the more commonly used Fisher’s exact test, e.g. [56], and both are presented in Figure 7.

**Figure 7:**
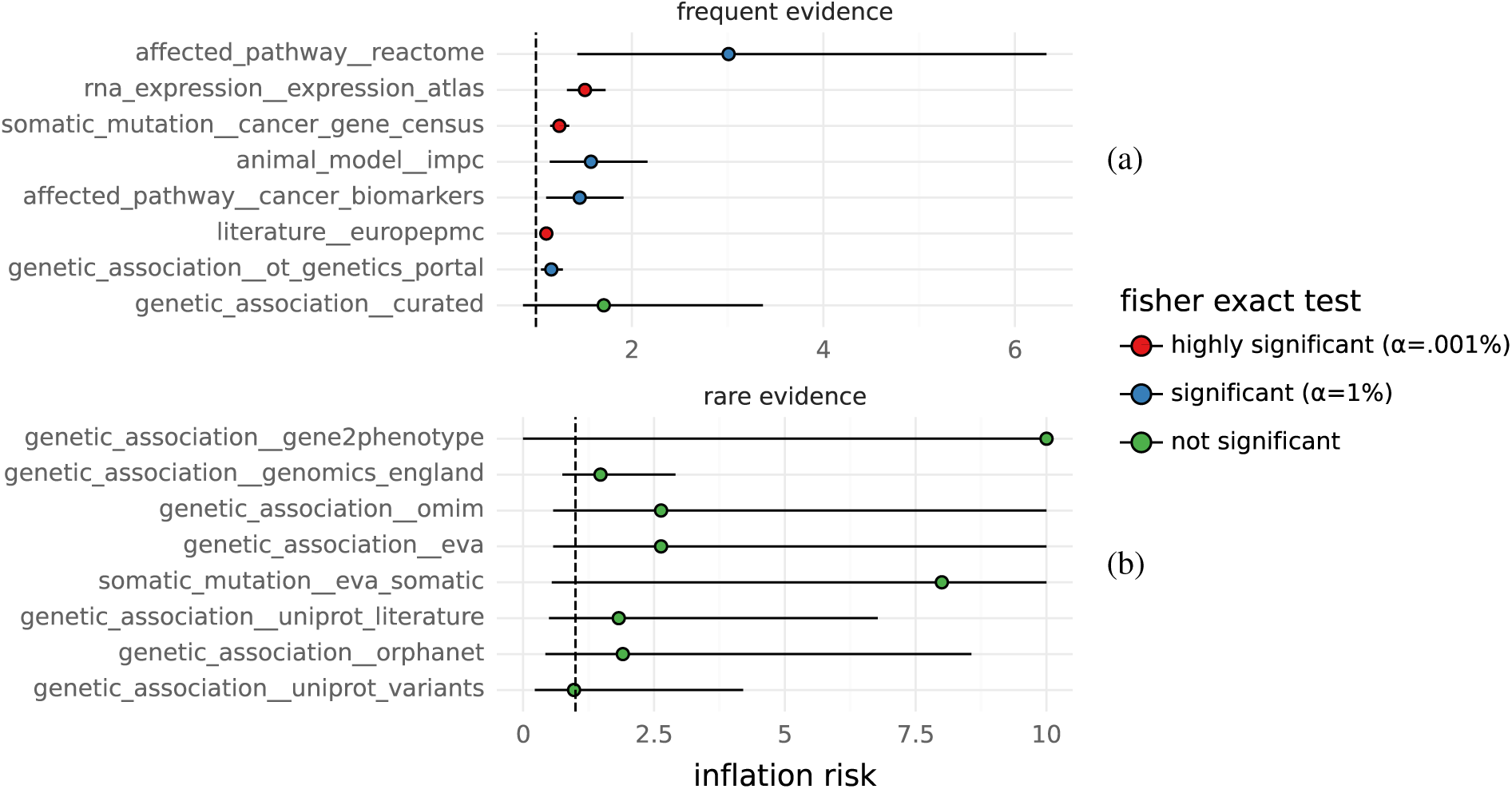
Inflation risk by feature. (a) Feature “inflation risk”, for more frequent features, reported as relative risk point estimates (dots) and Katz 99% confidence intervals (bars). These features are “frequent” in the sense that they have enough TD pairs where evidence emerged before **and** after a phase 2 transition to power a relatively narrow confidence interval. (b) Rare feature inflation risk with the same definition as and a wider range on the x-axis. All point estimates and confidence intervals are clamped to the range (0, 10) to minimize scale distortion from extreme estimates.

We find that evidence from Reactome is the worst offender by this metric, implying that it often only arises for TD pairs after a certain level of clinical success has been attained. We also find that long-running aggregators/curators of published research often focused on individual diseases/phenotypes, like Expression Atlas, IMPC, CGS and Cancer Biomarkers exhibit this form of inflation as well.

Sources of genetic evidence appear to be much less inflated, or have too little data to reach significance. This is to be expected for GWAS evidence arising from genome-wide, phenome-wide biobank consortia, however much of historical GWAS evidence is not phenome-wide. More context on how much this is likely to matter comes from [57] in which it was estimated that as few as 6% of 500 FDA-approved targets for non-cancer drugs arose from programs highly motivated by pre-existing genetic support and that “the remaining 94% were probably identified using conventional pharmacology, biochemistry or molecular biology approaches”. We then speculate that if the initiation of new drug programs was not historically motivated highly by the existence of genetic support, then the incentives for pursing new genetic evidence based on clinical and commercial success are likely to be minimized. This, in conjunction with existing precedent [7, 8, 6, 3, 9, 2] and our inflation results, ultimately led us to the use of genetic evidence without temporalization. In other words, we do not treat genetic evidence as longitudinal features like all others associated with TD pairs. A breakdown of which features are treated in which manner is provided in Table 1.

## 3 Conclusion

We have demonstrated that simple machine learning methods applied to longitudinal biomedical evidence from many sources can be used to identify combinations of drug targets and diseases that are 4-5x more likely to advance past phase 2 trials, and that this can be done without knowledge of molecular properties or trial design details. We have also shown that these methods are more precise in the extremes of their predictions than composite, heuristic scores like those from Open Targets. They also outperform such baselines by more comprehensive, traditional measures of classifier performance; however, we find this less compelling and easier to accomplish than improving performance among the upper tail of the opportunities implied by the very highest predictions. This framework would also support the addition of new lines of evidence over time well as it is designed to automatically determine the relevance of any new information without intervention. Lastly, we find that the space of present-day, undeveloped targets within a disease context that both exceed baseline levels of tractability and have a high predicted likelihood of clinical advancement is substantial. It is likely to grow quickly as well since the breadth of much of the underlying evidence is expanding rapidly [58, 59, 60].

## 4 Discussion

An important question, that remains difficult to answer, is how much target prioritization methods like the one explored in this study can improve the efficiency of drug discovery. One 2018 estimate suggests that improving success rates from phase 1 to approval by 1.4x (from 9.6% to 13.8%) would reduce the median R&D cost for a **single drug** by $480 million USD [61]. If a 2x increase in success rates from phase 1 to approval could be expected when pursuing only genetically validated TD pairs [7], then this would increase to a $686 million cost savings per drug. Pushing that even further to a 3x or 4x increase by incorporating more lines evidence, as we have done here, would suggest a per-drug cost savings of $1.03 billion and $1.37 billion, respectively. These savings would only be realized for drugs tested against targets and diseases with sufficient supporting evidence; however, we show in Section 2.6 that there are more than 8,000 such opportunities (i.e. untested, present-day TD pairs) where at least a 2.9x improvement in phase 2 success rates could be expected. This suggests that a substantial fraction of the 493 drug programs entering phase 2 trials each year [62] could potentially be replaced by programs from this pool.

Throughout the majority of this study, we focus on performance among the top 2% of TD pairs. Reasons for this are introduced in Section 2.1 and Section 2.3. It is constructive to expand on this choice by comparing it to alternatives, and noting that it is quite likely that many other predictors could improve performance more comprehensively. We offer some evidence of this through the use of a feature indicating how long a TD pair has been in phase 2 trials, which improves wholistic measures of ranking performance like ROC by 12 points (see Figure 4) and provides a particularly dramatic lift across the bottom 98% of TD pairs. While time is a necessary but not sufficient condition for clinical advancement of a TD pair, given that a single trial can take years to complete, it also correlates with how many distinct trials, drugs, companies, sponsors, investigators, etc. attempt to validate any one pair. We posit that this is crucial because even if a druggable, mechanistic link exists between a target and a disease, many (or even most) trials testing that link will fail (or not even start) for reasons unrelated to efficacy or safety of a drug [3]. Some of these reasons include 1) commercial factors like a lack of funding, pipeline reprioritization and competitive density, 2) administrative or logistical factors like a lack of enrollment, retention problems, poor trial design, drug supply chain shortages, epidemics (COVID-19) and 3) regulatory factors like legislative shifts or extensive regulatory approval delays. Many of these factors are fundamentally difficult or impossible to predict. Epidemic outbreaks, labor shortages leading to supply chain problems and regulatory shifts are examples of exogenous shocks for which accurate predictors are unlikely to exist. However, factors like funding, competition, enrollment and certain aspects of trial design have measurable predictors such as preceding venture capital and private equity investments, patent filings and revenue for related drugs, disease severity and prevalence, and the existence of biomarkers or other enabling factors for better trial designs (respectively). As a more detailed example, patient recruitment failures are the most common reason trials fail [3] and recruitment statistics are highly predictive of eventual trial outcomes [25, 24]. It stands to reason then that predicting trial enrollment failures well in advance of when a trial is even conceived, i.e. for a target-disease pair, should be possible because recruitment in trials is determined, in part, by how prevalent a disease is, how debilitating it is, what patient demographics it inflicts, whether existing treatments exist for it and how successful recruitment for past trials has been – all of which are measurable. Signals like this could be used to further improve performance since they better capture causal factors in non-biological advancement failures that time under development – the only non-biological feature in this study – does not.

Another notable focus of this study is on evidence for TD pairs that can be **directly** attributed to them. This is a departure from earlier research in this space like [7] that often includes measures of similarity between disease ontology terms to account for the fact that bridging disease nomenclatures used in clinical trial datasets with those used in databases maintaining genetic evidence is difficult. It is not uncommon for disease terms from either source to be sufficiently similar such that it is appropriate to consider them as equivalent. It can also be argued that similarity between disease terms offers an important dimension for expanding evidence in a biologically meaningful way, regardless of technical mapping issues. This is taken to further extremes in studies like [14] that propagates evidence using both measures of target similarity (e.g. using protein interaction networks) and measures of disease similarity (e.g. using ontologies and literature co-occurrence). We believe such expansion methods are very compatible with our approach by either using them to derive new predictors or to propagate predictions from models like these across networks or knowledge graphs.

All results we have presented are based on quasi-prospective outcomes. As described in Section 1, this means that the outcomes are not truly observed in the future relative to the information used to predict them. Both outcomes and predictors are assigned timestamps corresponding to publication or database dates that approximately capture when the events associated with them actually occurred. The accuracy of that approximation is difficult to determine, and publication dates can be particularly inaccurate when they lag evidence discovery by months or years. This does not pose a risk for information leaks in a quasi-prospective formulation though, as long as the date associated with an event does not precede its occurrence. Nevertheless, it is possible that publication dates are simply entered into a database incorrectly, clinical trial registries inaccurately represent when trials began or ended, or that information from future events ends up associated with the wrong targets and/or diseases. These are only a few possible risks for information leaks, and though each is relatively rare on its own, their combined impact could be substantial. And as with all forecasting models, the accuracy of future predictions is reliant on the extent to which the underlying data generating process may either change or deviate from anticipated changes. Fortunately, the evidence we employ as predictors here comes predominantly from molecular interactions in humans, animals or cells that are likely to remain stable in time, or shift on evolutionary time scales that eclipse drug development timelines. This summary on the subject from [14] is pertinent:

> It is important to bear in mind therefore that what we are measuring when looking at historical trial outcomes is not an unbiased measure of any given gene’s true disease associations, but rather a view on how useful a given evidence source or analytical method has been for choosing drug targets based on current and historical drug discovery practices. Dramatic changes in these practices in the future could render some of our conclusions obsolete, though the fundamental observation that genetic association itself is retained in molecular networks will remain valid.

Model performance evaluations based on prospective outcomes, as an alternative to quasi-prospective outcomes, offer a number of distinct advantages. For one, they mitigate nearly all the risks for information leaks mentioned previously by ensuring that all predictive information precedes clinical outcomes. They also offer a means to quantify those risks. An extension of this work might utilize the Open Targets database snapshots, dating back as far as 2016, to examine how much model performance differs between models trained on features from a snapshot and models trained on data from a current Open Targets release with features that are temporalized and then filtered to not exceed the year of the earlier snapshot. Similarly, that extension could examine how much model performance differs for a snapshot with and without longitudinal features by considering only outcomes that occurred after the snapshot. Such an approach could also be applied on a per-feature basis to gain more granular insight on which kinds of evidence pose the greatest risk. One clear disadvantage of prospective evaluation for this problem is that it requires years to accrue enough future clinical outcomes to be meaningful. This also means that predictors must omit that many years of accrued evidence as well, which can deflate performance compared to what is possible in the present day. Another disadvantage is that it is significantly more difficult to operationalize prospective evaluations, thereby implying that the decrease in risk of information leaks may be partially offset by an increase in technical risk.

## 5 Methods

All data used in this study comes from Open Targets with one minor exception. Open Targets provides PubMed identifiers as provenance for evidence without supplying publication dates or years in all cases. These dates are taken from [63] instead. Our primary results are based on the 23.12 Open Targets release, but we also use releases 23.09 and 23.06 as part of our sensitivity analysis in section 9.1. All code used is available at https://github.com/related-sciences/clinical_advancement_paper.

The features we derive from Open Targets come primarily from a single target-disease dataset containing only “direct” evidence [64]. These features are named according to the datasourceId field within this dataset, and the values for those features are defined as the cumulative, maximum score observed when ordering those scores by the years associated with the evidence underlying them. We also add 2 genetic features that are not immediately available. The first, genetic_association__curated is a union of all genetic association sources other than gene_burden and ot_genetics_portal. The second, genetic_association__omim, is defined as any EVA associations with a publication date.

The target-specific features we derive come from the “target prioritisation” dataset [32] in Open Targets. The only exception to this is a feature defining the maximum trial phase reached, which comes from the primary target-disease evidence dataset. Most scores in this dataset were not used because they cannot be temporalized. We make exceptions for those arising from large-scale, genome-wide assays like LOEUF from gnomAD [65] and tissue expression scores from Human Protein Atlas [66]. We retain a score derived from phenotype severity observed in mice knock-outs [67] as well. All three of these feature sources are unique in that they are imputed differently from all other features when absent in model training. The “closed-world” assumption discussed in Section 2.7 implies that a constant, zero imputation for most missing features is appropriate. This is not true for non-clinical, target-specific fields – they are mean-imputed instead.

All ridge regression models (e.g. RDG) are fit using the scikit-learn Ridge implementation and all gradient-boosted machine models (e.g. GBM-T) are fit using the LightGBM [68] algorithm. Ridge models are fit with very weak regularization (alpha=1e-6) and LightGBM models are fit with default settings. Variants of both models are fit, in some cases, with non-negative monotonicity constraints. An important caveat unique to our use of the ridge regression models is that when report results for feature ablations, as in Section 2.4.3, all features are actually used to train the models before using only the unablated features to make predictions.

Most results reported in this study consider only a subset of therapeutic areas within the Experimental Factor Ontology (EFO) [69]. All therapeutic areas are retained if they possessed at least 100 TD pairs with target-disease-specific evidence of any kind within our evaluation dataset. We also explicitly omit the “biological_process”, “pregnancy or perinatal disease”, “injury, poisoning or other complication”, “pregnancy or perinatal disease”, “medical procedure”, “infectious disease”, and “animal disease” therapeutic areas, as they are either not relevant or poorly represented. The “infectious disease” therapeutic area is a borderline case that we omit not because it is necessarily irrelevant, but because it has only 115 associated TD pairs. This makes it the most poorly represented therapeutic area that is possibly relevant. We remove it by explicit omission and use a 100 TD pair threshold rather than using a 115 TD pair threshold.

## 6 Limitations

There are a number of limitations and caveats with this study. These include:

1. Open Targets does not currently provide drug approvals with associated dates. This is a part of the reason why we have chosen to focus only on advancement beyond phase 2, and not phase 3 as well.
2. Not all Open Targets evidence can be associated with publication dates. This evidence is ignored when used to construct longitudinal predictors; however, this limitation does not apply to Section 2.6.
3. The rankings of undeveloped TD pairs in Section 2.6 are presented directly from Open Targets with no additional curation or vetting. We recognize that some of these TD pairs are for uninformative disease terms, e.g. the association between “CTLA4” and “autoimmune disease” in Supplementary Figure 15, and we opt not address these cases to best reflect the state of rankings within Open Targets. Several more computational steps are necessary in practice to identify a more enriched set of actionable TD pairs.
4. The Open Targets composite score we use (OTS) is not equivalent to the overall composite score for TD pairs provided by Open Targets [70]. The score they provide is not longitudinal. We create an approximation of it over time based on the same weighting scheme across evidence types, but without a harmonic sum aggregation function. Maximum scores for the same target, disease, year and evidence type combination are used instead.
5. Estimating an absolute number of TD pairs associated with any criteria is difficult. In Section 2.6, we present absolute counts of undeveloped TD pairs with tractable targets and a high likelihood of clinical success. These counts may be inflated by the fact that the associated targets and diseases are so sufficiently similar that they ought not to be counted as distinct examples. Defining a threshold for similarity to combat this problem is difficult and application-specific though, so we do not offer any solutions here.
6. The source of OMIM associations we use is derived from a subset of associations available in EVA. This subset is defined by associations with a linked publication. This is only an approximation, but our internal research shows that 98% of EVA records with linked publications also have at least one OMIM submission.

## 7 Data Availability

All code, data and analysis used for this study, when not noted as proprietary, is available at https://github.com/related-sciences/clinical_advancement_paper.

## 8 Appendix

**Figure 8:**
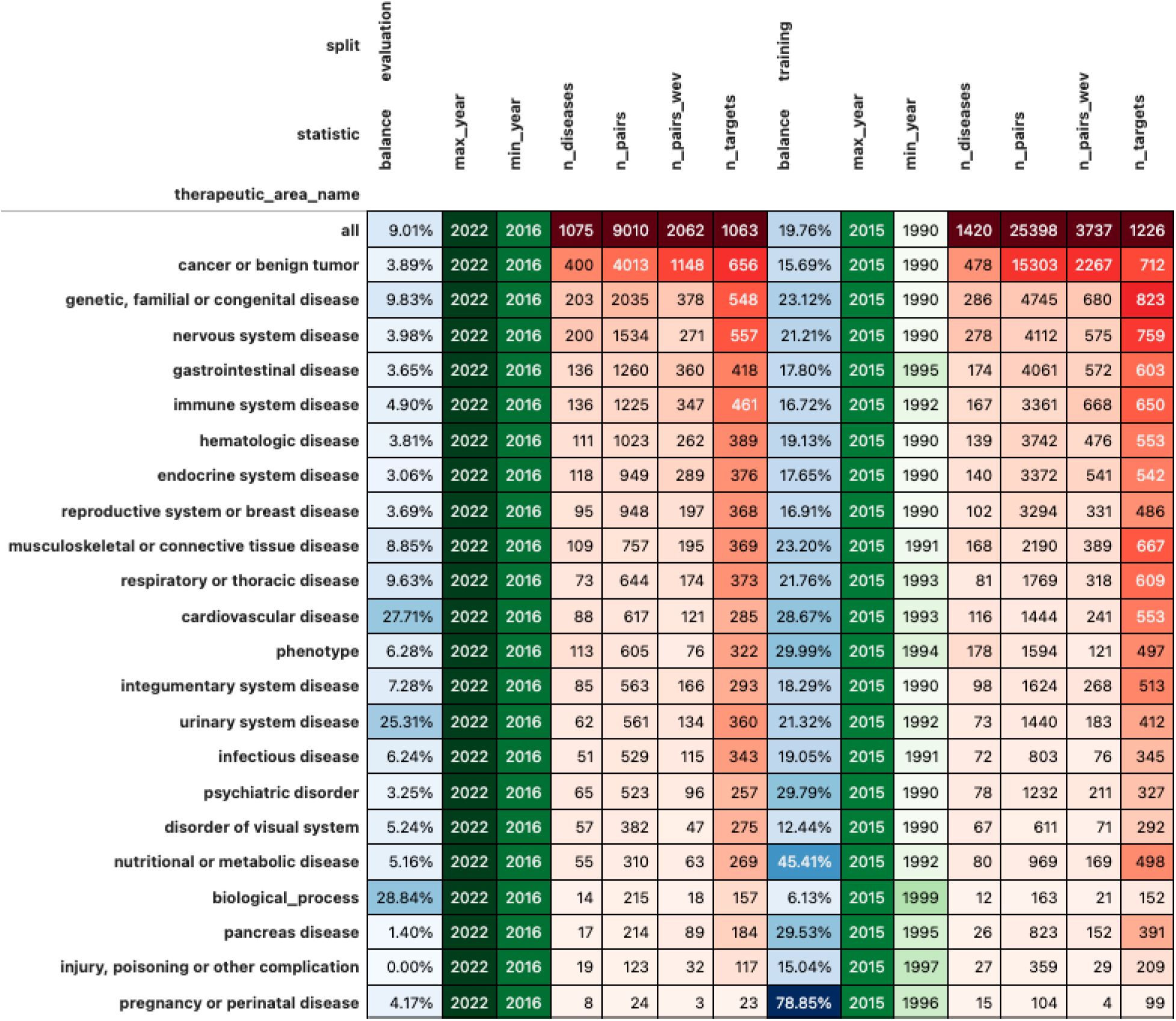
Training and evaluation dataset summary statistics. balance: percentage of TD pairs that eventually advanced beyond phase 2 min_year: earliest year in which a TD pair has first advanced to phase 2 n_targets: number of unique targets n_diseases: number of unique diseases n_pairs: number of TD pairs (equal to number of records) n_pairs_wev: number of TD pairs with directly associated evidence (i.e. not just target or disease evidence).

**Table 1:** Features used in modeling and analysis.

|  | feature | entity | kind |
| --- | --- | --- | --- |
| 1 | disease__clinical__phase_max__reached | disease | temporal |
| 2 | target__clinical__phase_max__reached | target | temporal |
| 3 | target__genetic_constraint | target | static |
| 4 | target__mouse_ko_score | target | static |
| 5 | target__tissue_distribution | target | static |
| 6 | target__tissue_specificity | target | static |
| 7 | target_disease__affected_pathway__cancer_biomarkers | target_disease | temporal |
| 8 | target_disease__affected_pathway__crispr | target_disease | temporal |
| 9 | target_disease__affected_pathway__crispr_screen | target_disease | temporal |
| 10 | target_disease__affected_pathway__progeny | target_disease | temporal |
| 11 | target_disease__affected_pathway__reactome | target_disease | temporal |
| 12 | target_disease__affected_pathway__slapenrich | target_disease | temporal |
| 13 | target_disease__affected_pathway__sysbio | target_disease | temporal |
| 14 | target_disease__animal_model__impc | target_disease | temporal |
| 15 | target_disease__genetic_association__clingen | target_disease | static |
| 16 | target_disease__genetic_association__curated | target_disease | static |
| 17 | target_disease__genetic_association__eva | target_disease | static |
| 18 | target_disease__genetic_association__gene2phenotype | target_disease | static |
| 19 | target_disease__genetic_association__gene_burden | target_disease | static |
| 20 | target_disease__genetic_association__genomics_england | target_disease | static |
| 21 | target_disease__genetic_association__omim | target_disease | static |
| 22 | target_disease__genetic_association__orphanet | target_disease | static |
| 23 | target_disease__genetic_association__ot_genetics_portal | target_disease | static |
| 24 | target_disease__genetic_association__uniprot_literature | target_disease | static |
| 25 | target_disease__genetic_association__uniprot_variants | target_disease | static |
| 26 | target_disease__known_drug__chembl | target_disease | temporal |
| 27 | target_disease__literature__europepmc | target_disease | temporal |
| 28 | target_disease__outcome__advanced | target_disease | temporal |
| 29 | target_disease__rna_expression__expression_atlas | target_disease | temporal |
| 30 | target_disease__somatic_mutation__cancer_gene_census | target_disease | temporal |
| 31 | target_disease__somatic_mutation__eva_somatic | target_disease | temporal |
| 32 | target_disease__somatic_mutation__intogen | target_disease | temporal |
| 33 | target_disease__time__transition | target_disease | temporal |

**Table 2:** Tractability bucket assignments.

|  | evidence | modality | confidence |
| --- | --- | --- | --- |
| 1 | Phase 1 Clinical | OC | LOW |
| 2 | Advanced Clinical | OC | MED |
| 3 | Approved Drug | OC | HIGH |
| 4 | GO CC med conf | AB | LOW |
| 5 | Human Protein Atlas loc | AB | LOW |
| 6 | UniProt SigP or TMHMM | AB | LOW |
| 7 | UniProt loc med conf | AB | LOW |
| 8 | GO CC high conf | AB | MED |
| 9 | UniProt loc high conf | AB | MED |
| 10 | Advanced Clinical | AB | HIGH |
| 11 | Approved Drug | AB | HIGH |
| 12 | Phase 1 Clinical | AB | HIGH |
| 13 | Database Ubiquitination | PR | LOW |
| 14 | Half-life Data | PR | LOW |
| 15 | Small Molecule Binder | PR | LOW |
| 16 | Literature | PR | MED |
| 17 | UniProt Ubiquitination | PR | MED |
| 18 | Advanced Clinical | PR | HIGH |
| 19 | Phase 1 Clinical | PR | HIGH |
| 20 | Druggable Family | SM | LOW |
| 21 | High-Quality Pocket | SM | LOW |
| 22 | Med-Quality Pocket | SM | LOW |
| 23 | High-Quality Ligand | SM | MED |
| 24 | Structure with Ligand | SM | MED |
| 25 | Advanced Clinical | SM | HIGH |
| 26 | Approved Drug | SM | HIGH |
| 27 | Phase 1 Clinical | SM | HIGH |

## 9 Supplementary Material

**Figure 9:**
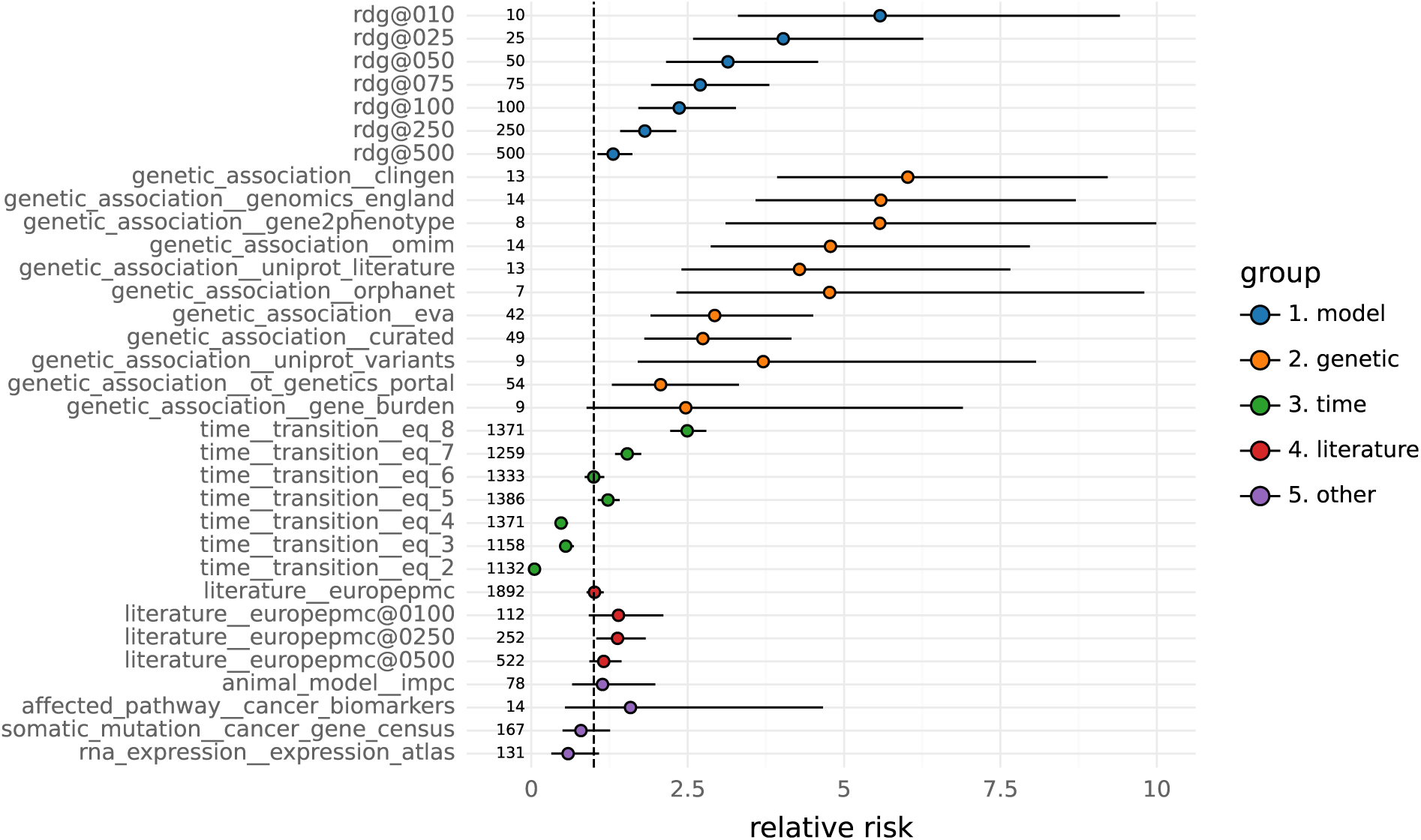
Performance of individual features and predictive scores as measured by relative risk. RR estimates are shown as dots with bars indicating 90% Katz confidence intervals. RDG model results denoted by rdg@N indicate performance for the N top TD pairs. The same convention is used for literature evidence and the time_transition_eq_X convention denotes RR estimates when the time since the phase 2 transition is equal to X years. The omim, eva, and ot_genetics_portal features correspond to the OMIM, EVA and OTG baselines of Figure 3, respectively. All other features are assessed based on their existence. The counts along the origin indicate how many TD pairs were used to compute the RR numerator.

**Figure 10:**
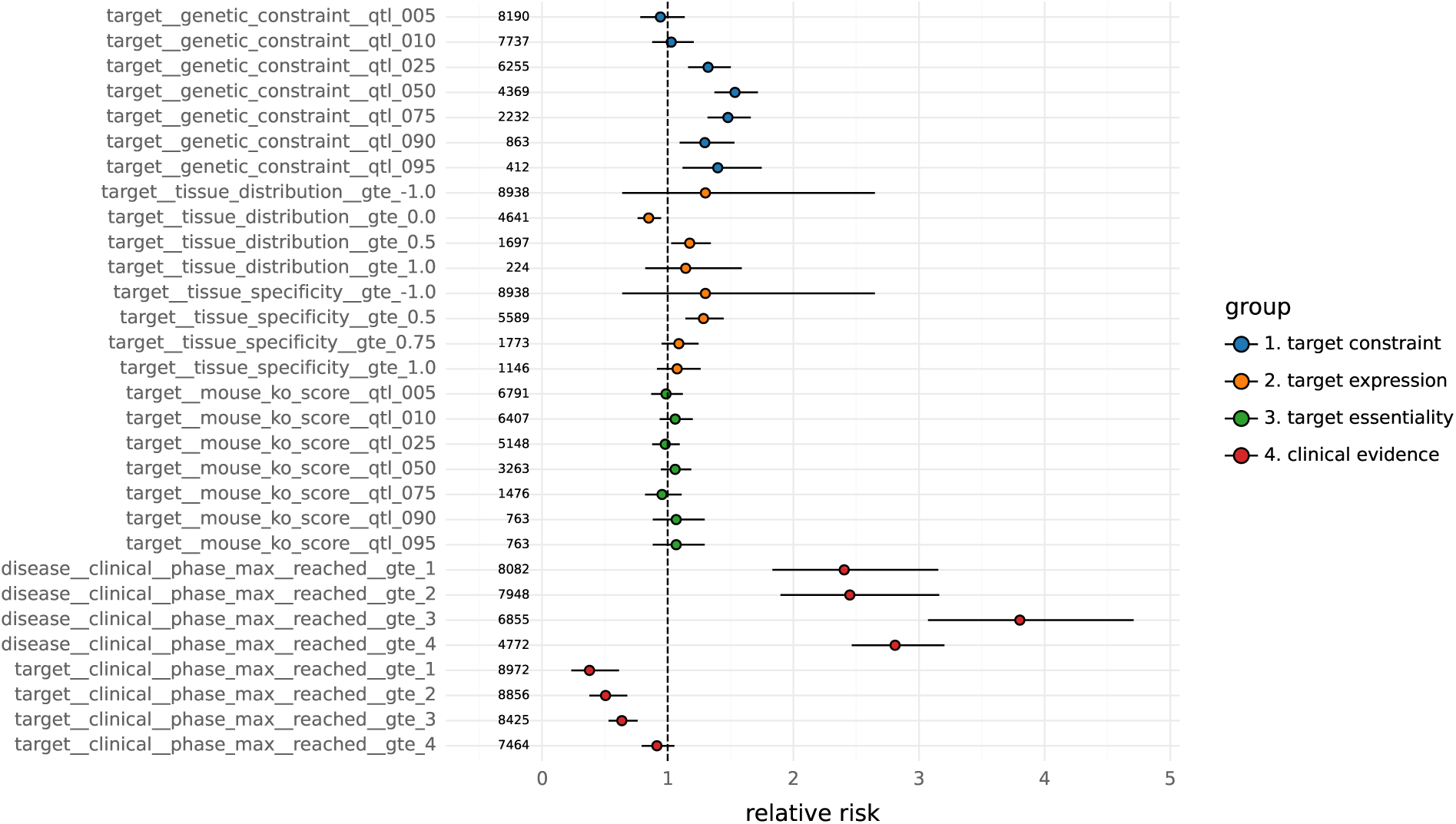
Relative risk scores for target-only and disease-only features. RR estimates are shown as dots with bars indicating 90% Katz confidence intervals. The features ending with qtl_Q denote binary indicators constructed from cases where the feature meets or exceeds quantile Q of its distribution. The features ending with gte_X denote indicators for when the feature meets or exceeds a specific value X. The counts along the origin indicate how many TD pairs were used to compute the RR numerator.

**Figure 11:**
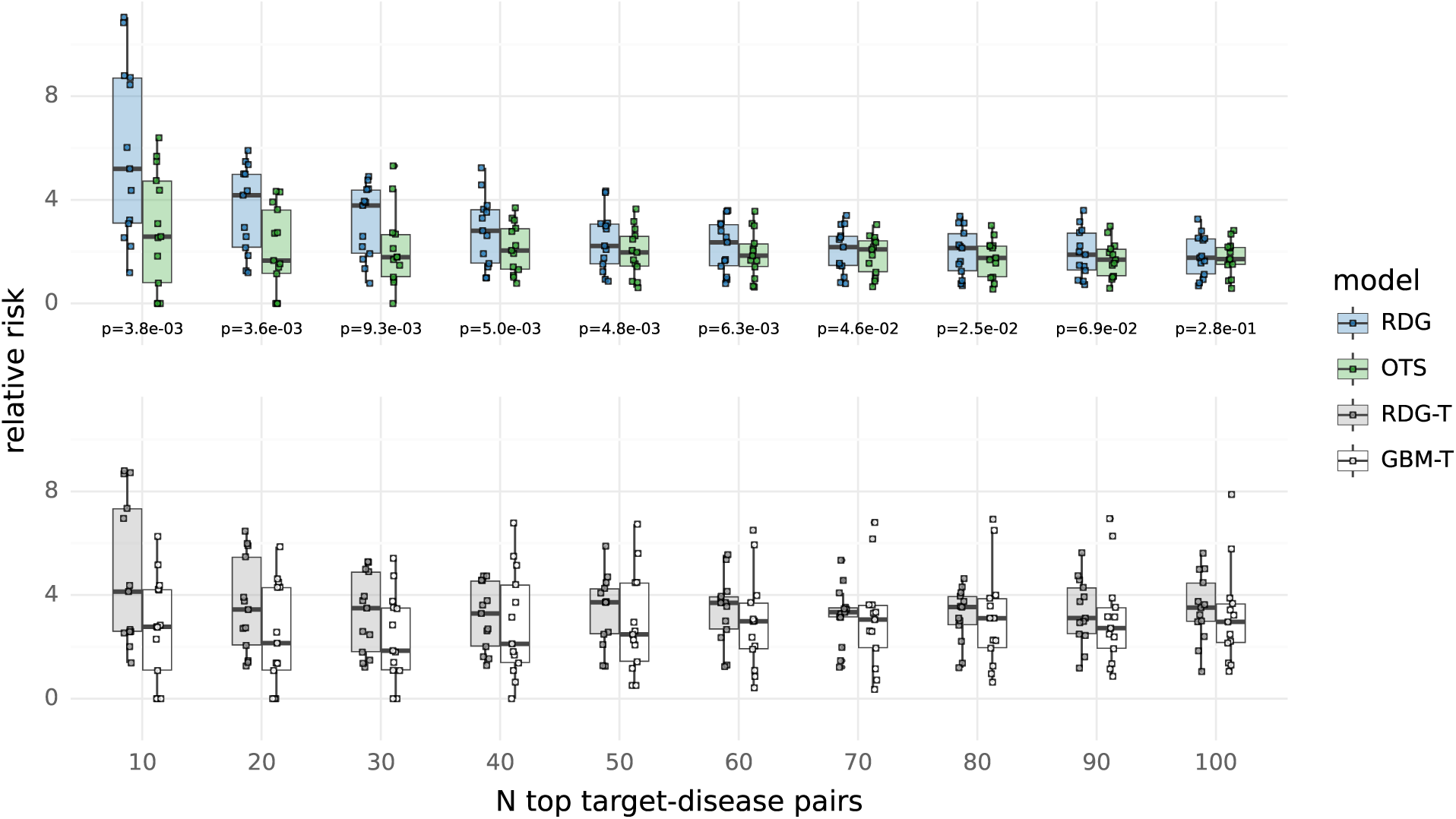
Relative risk distributions across select therapeutic areas. Relative risk point estimates for each of 13 therapeutic areas are shown as dots with boxplots underneath summarizing their distribution. P-values on the bottom margin of the top facet are computed from a one-sided Wilcoxon signed-rank test with the alternative that the RDG model RR average across therapeutic areas exceeds the OTS average at a given number of top ranking TD pairs (on the x-axis).

**Figure 12:**
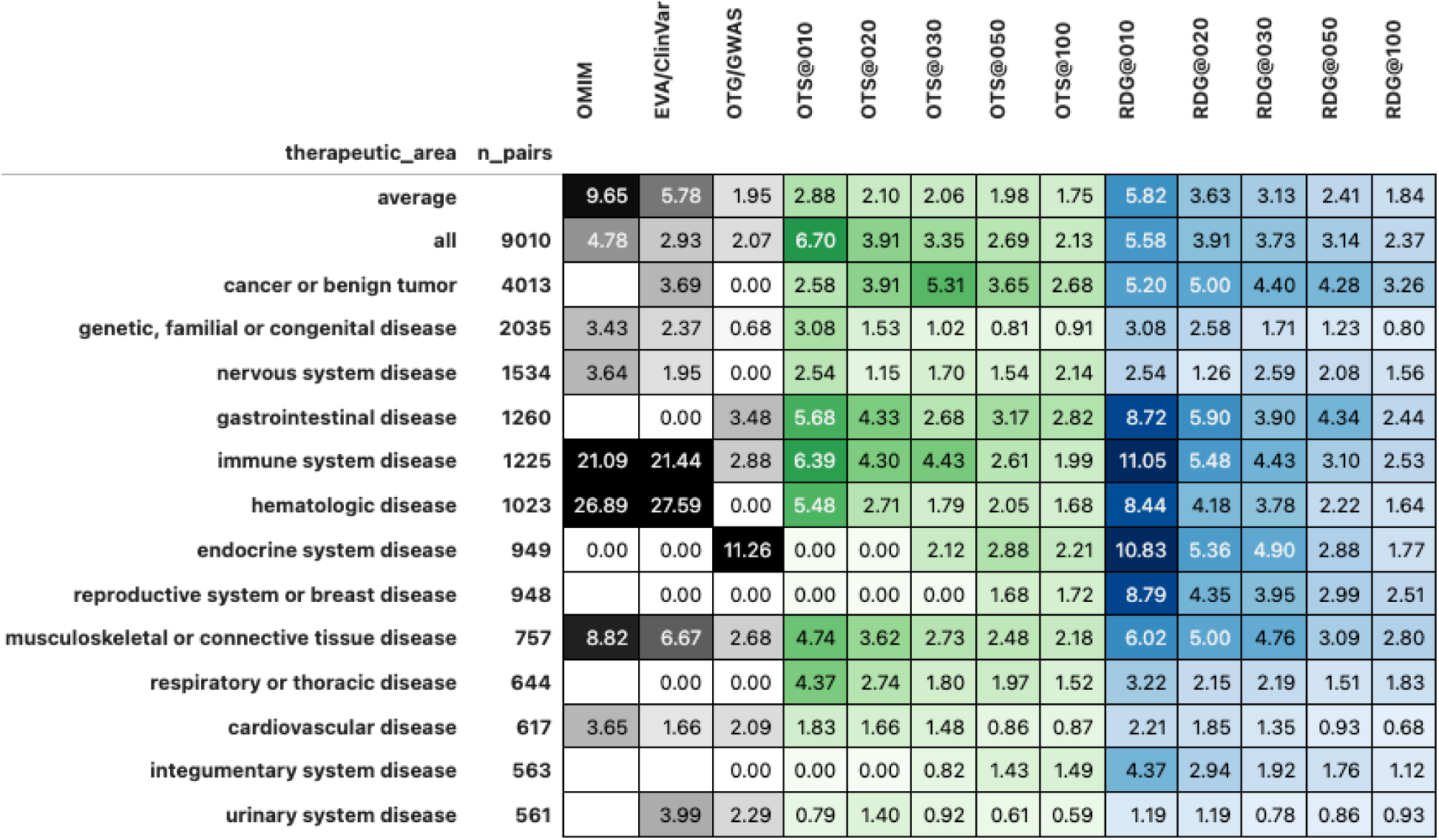
Relative risk scores by method, benchmark and therapeutic area. The average therapeutic area denotes mean values across all other therapeutic areas except for all, which is an ungrouped estimate across all diseases regardless of therapeutic area. The n_pairs field indicates the number of TD pairs used to compute the RR estimate in each cell. The OTS@N and RDG@N columns indicate RR estimates for the top N TD pairs from the OTS and RDG methods, respectively.

**Figure 13:**
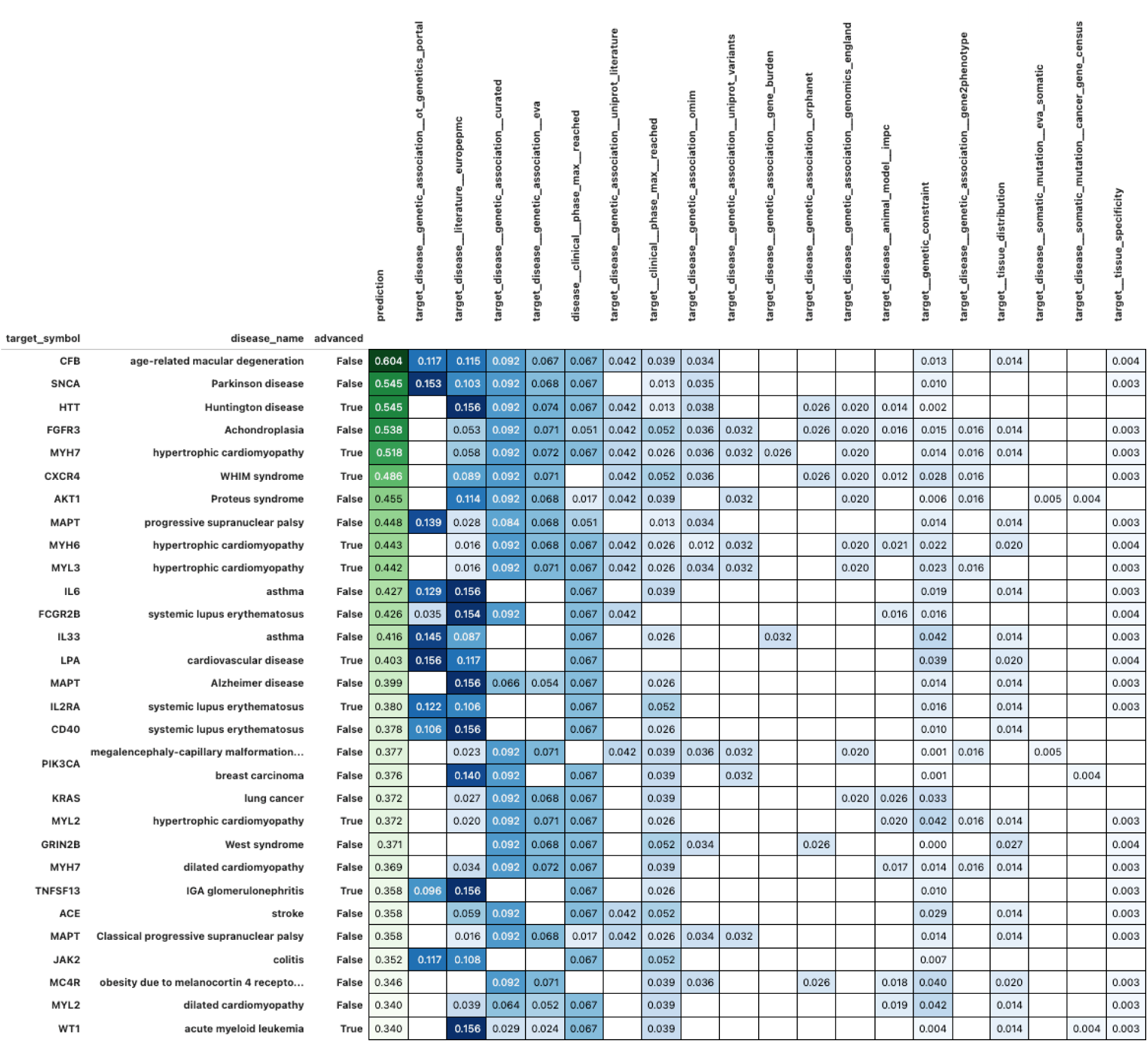
Top RDG model evaluation dataset predictions. Feature contributions are shown as the product of their underlying values and the RDG coefficients. The advanced field indicates whether the associated TD pair advanced beyond phase 2 as of 2024. The prediction field shows the final RDG model prediction for each case.

**Figure 14:**
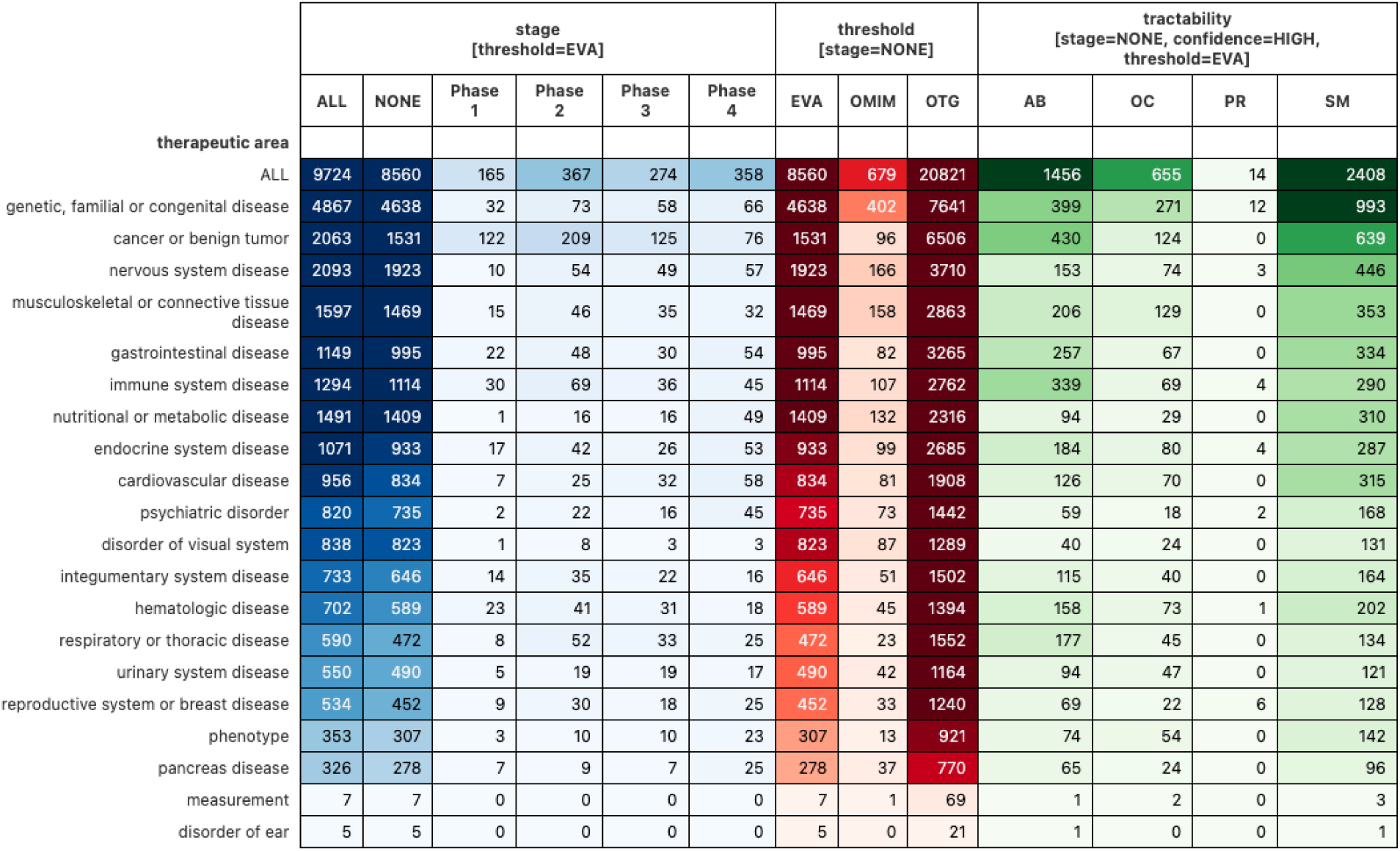
Present-day target-disease pair counts by stage, likelihood of advancement and tractability. The stage panel contains counts by maximum trial phase reached, the threshold panel contains counts of pairs with a RDG model score exceeding that of the associated benchmark for only undeveloped pairs, and the tractability panel shows pair frequencies among undeveloped pairs exceeding the EVA threshold that also have a HIGH tractability rating as defined in Supplementary Table 2. See Figure 6 for more details and a more detailed visual summary of these same results for the ALL therapeutic area, which simply indicates counts across all diseases regardless of therapeutic area.

**Figure 15:**
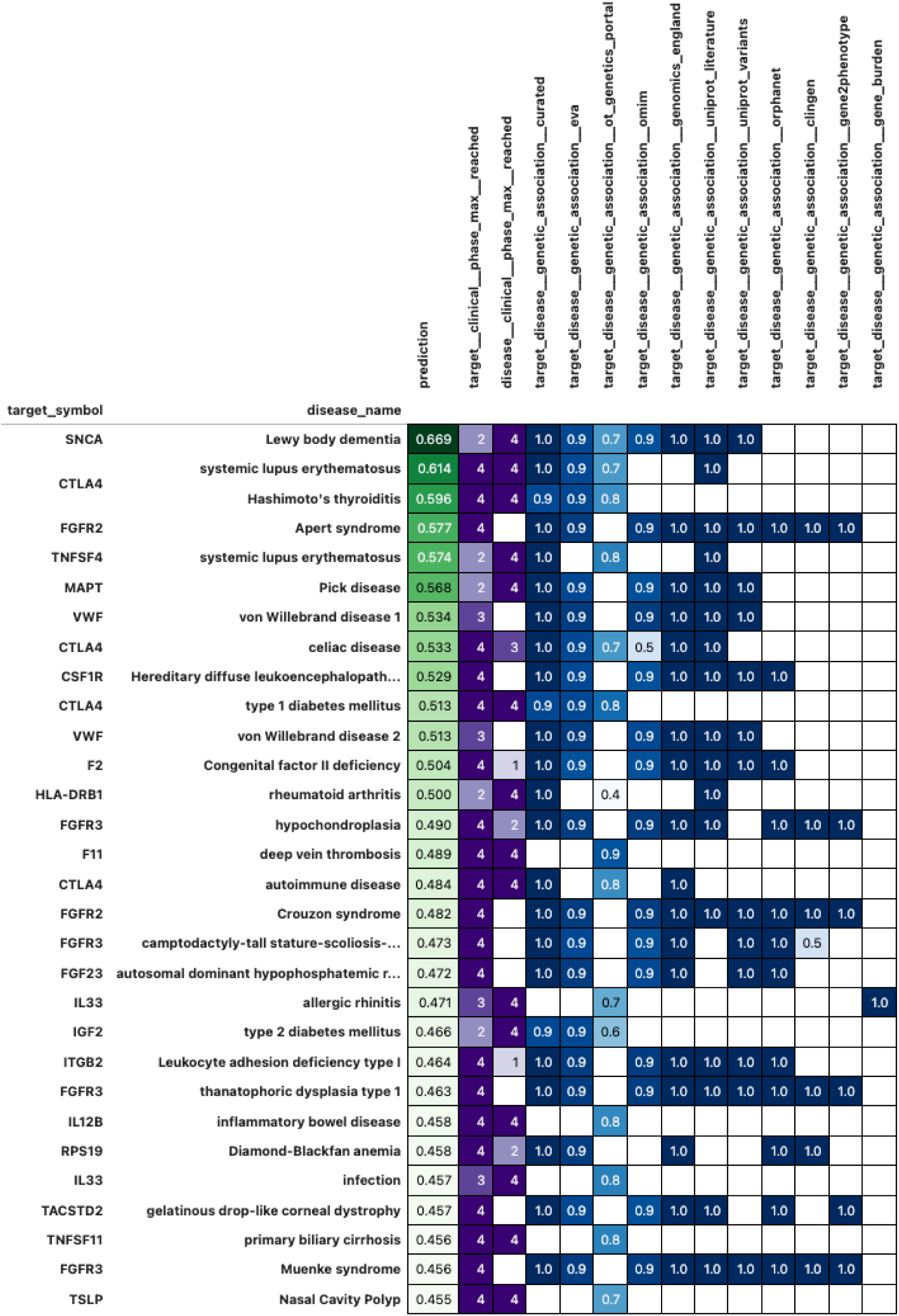
Top ranked undeveloped, antibody-enabled target-disease pairs. Present-day, undeveloped, antibody-enabled TD pairs with the highest RDG model predictions (shown in the prediction column) are presented here. These pairs are “undeveloped” in the sense that they have no known clinical trials. Furthermore, they are “antibody-enabled” in the sense that the targets for each pair have been previously subjected to antibody-based drugs in clinical trials. More details and context are provided in Figure 6.

### 9.1 Sensitivity

In order to validate the stability of our findings in Section 2.4, we repeat this analysis across 18 different configurations listed in Supplementary Table 3. This includes 3 separate versions of Open Targets, 3 choices for the year defining the split between training and evaluation data and 2 choices for the length of the minimum advancement window (in years).

We find that the mean RR values from the RDG model consistently exceed the OTS model in all configurations among the very highest rankings (N=10) and also exceed the OTS model in all configurations except for 1 for N between 20 and 60. This data is shown in Supplementary Figure 16. The significance of these differences drops notably after N=40, which can be seen in the distribution of p-values from a Wilcoxon signed-rank test shown in Supplementary Figure 17.

**Table 3:**
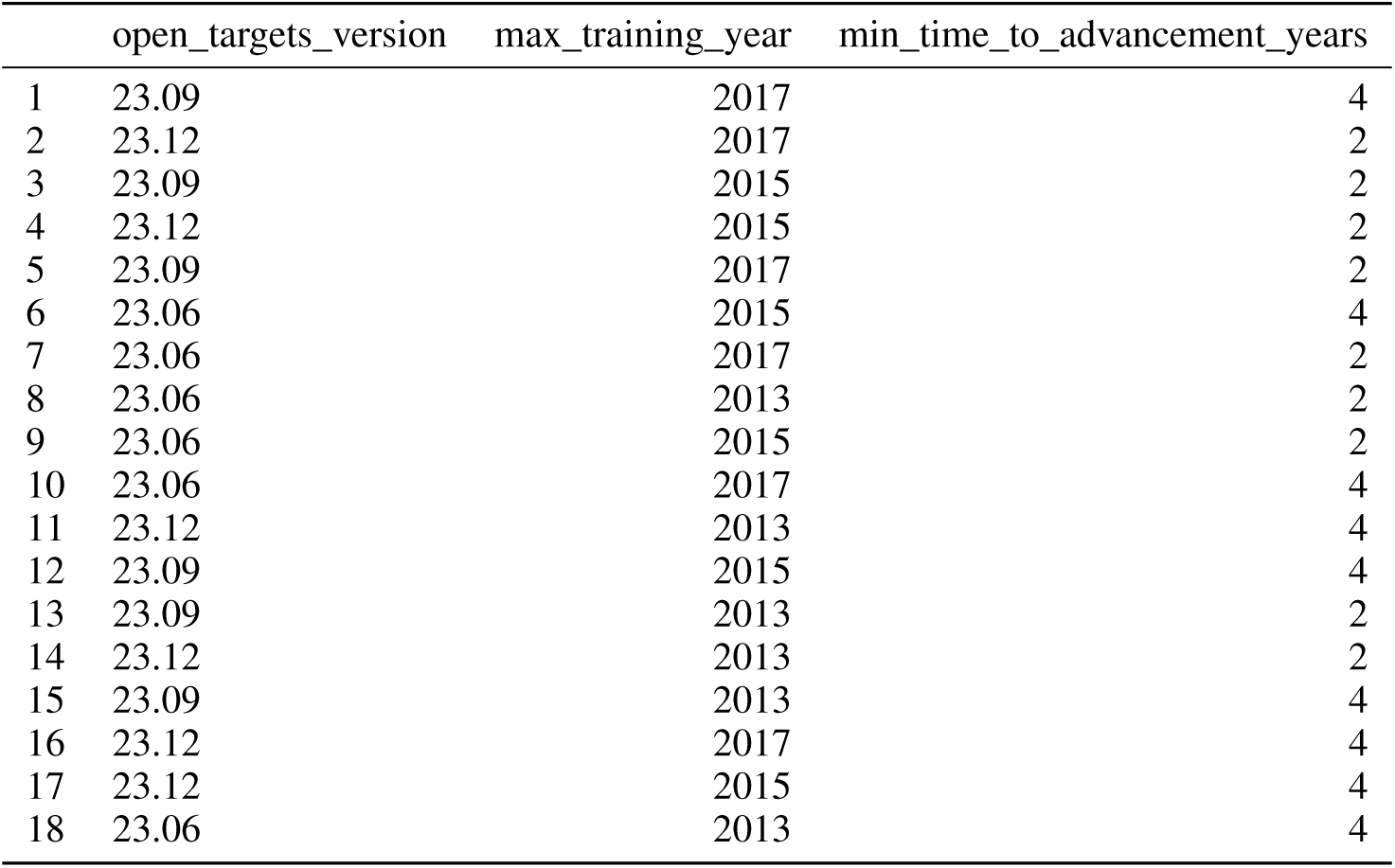
Configurations for sensitivity analysis.

**Figure 16:**
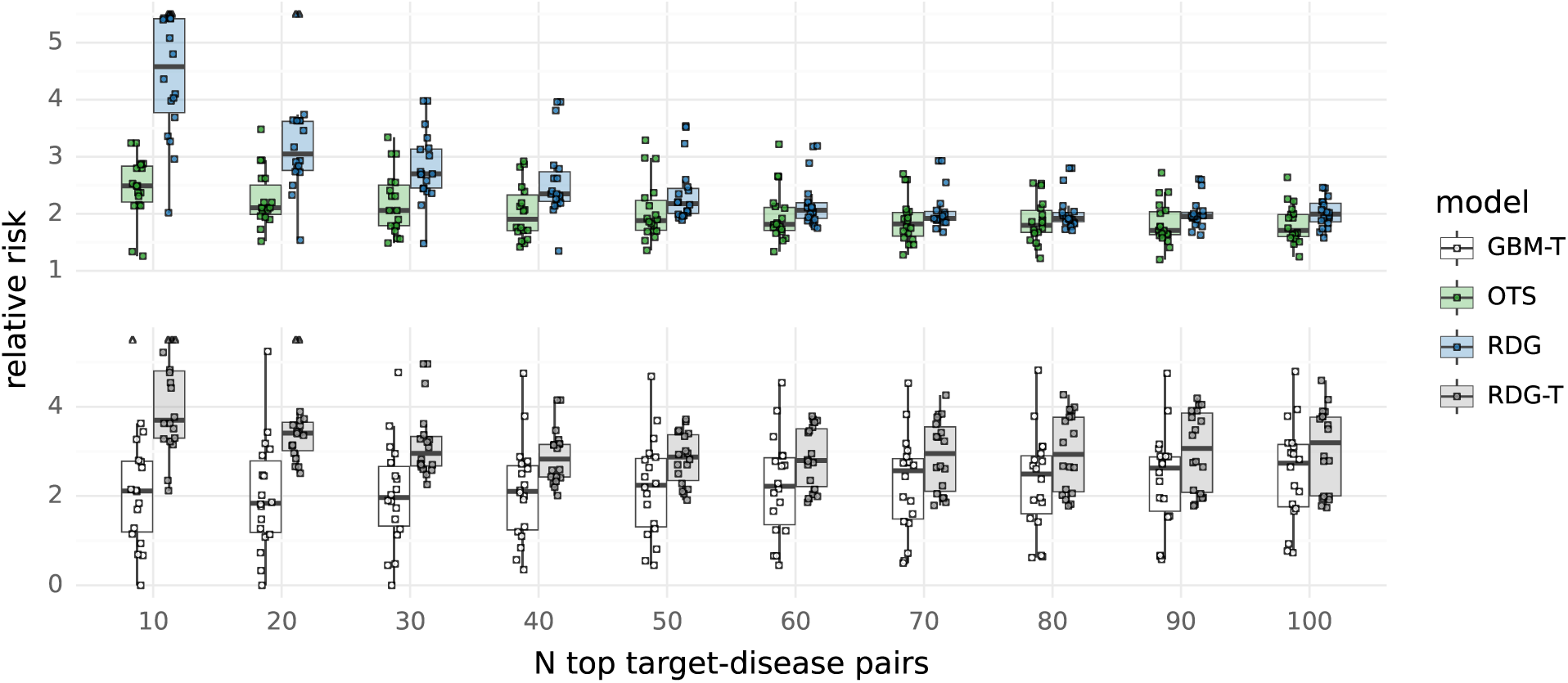
Relative risk distributions across configurations in sensitivity analysis. The distribution of the mean RR values displayed for a single configuration in Figure 2 is shown here across 18 configurations.

**Figure 17:**
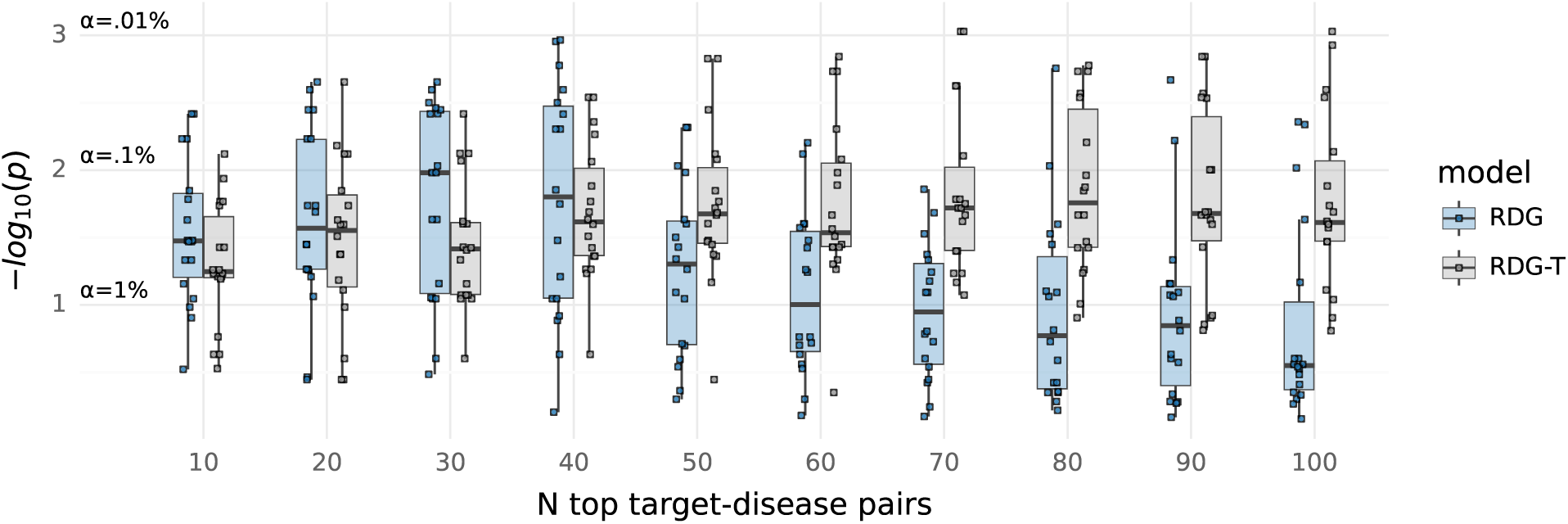
P-value distributions across configurations in sensitivity analysis. The distribution of the p-values displayed for a single configuration in Supplementary Figure 11 is shown here across 18 configurations.

## References

[1] David Cook, Dearg Brown, Robert Alexander, Ruth March, Paul Morgan, Gemma Satterthwaite, and Menelas N Pangalos. Lessons learned from the fate of astrazeneca’s drug pipeline: a five-dimensional framework. Nature reviews. Drug discovery, 13(6):419—431, June 2014.

[2] David Ochoa, Mohd Karim, Maya Ghoussaini, David G Hulcoop, Ellen M McDonagh, and Ian Dunham. Human genetics evidence supports two-thirds of the 2021 fda-approved drugs. Nature reviews. Drug discovery, 21(8):551, August 2022.

[3] Olesya Razuvayevskaya, Irene Lopez, Ian Dunham, and David Ochoa. Why clinical trials stop: The role of genetics. medRxiv, 2023.

[4] Polina V Rusina, Maria J Falaguera, Juan Maria R Romero, Ellen M McDonagh, Ian Dunham, and David Ochoa. Genetic support for fda-approved drugs over the past decade. Nature reviews. Drug discovery, 22(11):864, November 2023.

[5] Maya Ghoussaini, Matthew R Nelson, and Ian Dunham. Future prospects for human genetics and genomics in drug discovery. Current opinion in structural biology, 80:102568, June 2023.

[6] Eric Vallabh Minikel, Jeffery L Painter, Coco Chengliang Dong, and Matthew R. Nelson. Refining the impact of genetic evidence on clinical success. medRxiv, 2023.

[7] Matthew R Nelson, Hannah Tipney, Jeffery L Painter, Judong Shen, Paola Nicoletti, Yufeng Shen, Aris Floratos, Pak Chung Sham, Mulin Jun Li, Junwen Wang, Lon R Cardon, John C Whittaker, and Philippe Sanseau. The support of human genetic evidence for approved drug indications. Nat. Genet., 47(8):856–860, August 2015.

[8] Emily A King, J Wade Davis, and Jacob F Degner. Are drug targets with genetic support twice as likely to be approved? revised estimates of the impact of genetic support for drug mechanisms on the probability of drug approval. PLoS Genet., 15(12):e1008489, December 2019.

[9] Ben Kinnersley, Amit Sud, Elizabeth A Coker, Joseph E Tym, Patrizio Di Micco, Bissan Al-Lazikani, and Richard S Houlston. Leveraging human genetics to guide cancer drug development. JCO clinical cancer informatics, 2:1—11, December 2018.

[10] Marie C Sadler, Chiara Auwerx, Patrick Deelen, and Zoltán Kutalik. Multi-layered genetic approaches to identify approved drug targets. Cell Genom., 3(7):100341, July 2023.

[11] Áine Duffy, Ben Omega Petrazzini, David Stein, Joshua K Park, Iain S Forrest, Kyle Gibson, Ha My Vy, Robert Chen, Carla Márquez-Luna, Matthew Mort, Marie Verbanck, Avner Schlessinger, Yuval Itan, David N Cooper, Ghislain Rocheleau, Daniel M Jordan, and Ron Do. Development of a human genetics-guided priority score for 19,365 genes and 399 drug indications. Nature genetics, 56(1):51—59, January 2024.

[12] Gautier Koscielny, Peter An, Denise Carvalho-Silva, Jennifer A Cham, Luca Fumis, Rippa Gasparyan, Samiul Hasan, Nikiforos Karamanis, Michael Maguire, Eliseo Papa, Andrea Pierleoni, Miguel Pignatelli, Theo Platt, Francis Rowland, Priyanka Wankar, A Patrícia Bento, Tony Burdett, Antonio Fabregat, Simon Forbes, Anna Gaulton, Cristina Yenyxe Gonzalez, Henning Hermjakob, Anne Hersey, Steven Jupe, Ş enay Kafkas, Maria Keays, Catherine Leroy, Francisco-Javier Lopez, Maria Paula Magarinos, James Malone, Johanna McEntyre, Alfonso Munoz-Pomer Fuentes, Claire O’Donovan, Irene Papatheodorou, Helen Parkinson, Barbara Palka, Justin Paschall, Robert Petryszak, Naruemon Pratanwanich, Sirarat Sarntivijal, Gary Saunders, Konstantinos Sidiropoulos, Thomas Smith, Zbyslaw Sondka, Oliver Stegle, Y Amy Tang, Edward Turner, Brendan Vaughan, Olga Vrousgou, Xavier Watkins, Maria-Jesus Martin, Philippe Sanseau, Jessica Vamathevan, Ewan Birney, Jeffrey Barrett, and Ian Dunham. Open targets: a platform for therapeutic target identification and validation. Nucleic Acids Res., 45(D1):D985–D994, January 2017.

[13] Hai Fang, ULTRA-DD Consortium, Hans De Wolf, Bogdan Knezevic, Katie L Burnham, Julie Osgood, Anna Sanniti, Alicia Lledó Lara, Silva Kasela, Stephane De Cesco, Jörg K Wegner, Lahiru Handunnetthi, Fiona E McCann, Liye Chen, Takuya Sekine, Paul E Brennan, Brian D Marsden, David Damerell, Chris A O’Callaghan, Chas Bountra, Paul Bowness, Yvonne Sundström, Lili Milani, Louise Berg, Hinrich W Göhlmann, Pieter J Peeters, Benjamin P Fairfax, Michael Sundström, and Julian C Knight. A genetics-led approach defines the drug target landscape of 30 immune-related traits. Nature genetics, 51(7):1082—1091, July 2019.

[14] Aidan MacNamara, Nikolina Nakic, Ali Amin Al Olama, Cong Guo, Karsten B Sieber, Mark R Hurle, and Alex Gutteridge. Network and pathway expansion of genetic disease associations identifies successful drug targets. Scientific reports, 10(1):20970, December 2020.

[15] Chaohui Bao, Hengru Wang, and Hai Fang. Genomic evidence supports the recognition of endometriosis as an inflammatory systemic disease and reveals disease-specific therapeutic potentials of targeting neutrophil degranulation. Front. Immunol., 13:758440, March 2022.

[16] Inigo Barrio-Hernandez and Pedro Beltrao. Network analysis of genome-wide association studies for drug target prioritisation. Current opinion in chemical biology, 71:102206, December 2022.

[17] Inigo Barrio-Hernandez, Jeremy Schwartzentruber, Anjali Shrivastava, Noemi Del-Toro, Asier Gonzalez, Qian Zhang, Edward Mountjoy, Daniel Suveges, David Ochoa, Maya Ghoussaini, Glyn Bradley, Henning Hermjakob, Sandra Orchard, Ian Dunham, Carl A Anderson, Pablo Porras, and Pedro Beltrao. Network expansion of genetic associations defines a pleiotropy map of human cell biology. Nature genetics, 55(3):389—398, March 2023.

[18] Saee Paliwal, Alex de Giorgio, Daniel Neil, Jean-Baptiste Michel, and Alix Mb Lacoste. Preclinical validation of therapeutic targets predicted by tensor factorization on heterogeneous graphs. Sci. Rep., 10(1):18250, October 2020.

[19] Camilo Ruiz, Marinka Zitnik, and Jure Leskovec. Identification of disease treatment mechanisms through the multiscale interactome. Nature communications, 12(1):1796, March 2021.

[20] Srivamshi Pittala, William Koehler, Jonathan Deans, Daniel Salinas, Martin Bringmann, Katharina Sophia Volz, and Berk Kapicioglu. Relation-weighted link prediction for disease gene identification, 2020.

[21] Ozlem Muslu, Charles Tapley Hoyt, Mauricio Lacerda, Martin Hofmann-Apitius, and Holger Frohlich. Guiltytar-gets: Prioritization of novel therapeutic targets with network representation learning. IEEE/ACM transactions on computational biology and bioinformatics, 19(1):491—500, 2022.

[22] Petrina Kamya, Ivan V Ozerov, Frank W Pun, Kyle Tretina, Tatyana Fokina, Shan Chen, Vladimir Naumov, Xi Long, Sha Lin, Mikhail Korzinkin, Daniil Polykovskiy, Alex Aliper, Feng Ren, and Alex Zhavoronkov. Pandaomics: An ai-driven platform for therapeutic target and biomarker discovery. Journal of chemical information and modeling, February 2024.

[23] Alex Aliper, Roman Kudrin, Daniil Polykovskiy, Petrina Kamya, Elena Tutubalina, Shan Chen, Feng Ren, and Alex Zhavoronkov. Prediction of clinical trials outcomes based on target choice and clinical trial design with multi-modal artificial intelligence. Clinical pharmacology and therapeutics, 114(5):972—980, November 2023.

[24] Kien Wei Siah, Nicholas W Kelley, Steffen Ballerstedt, Björn Holzhauer, Tianmeng Lyu, David Mettler, Sophie Sun, Simon Wandel, Yang Zhong, Bin Zhou, Shifeng Pan, Yingyao Zhou, and Andrew W Lo. Predicting drug approvals: The novartis data science and artificial intelligence challenge. Patterns (New York, N.Y.), 2(8):100312, August 2021.

[25] Andrew W. Lo, Kien Wei Siah, and Chi Heem Wong. Machine Learning With Statistical Imputation for Predicting Drug Approvals. Harvard Data Science Review, 1(1), jul 1 2019. https://hdsr.mitpress.mit.edu/pub/ct67j043.

[26] David Narganes-Carlón, Daniel J Crowther, and Ewan R Pearson. A publication-wide association study (pwas), historical language models to prioritise novel therapeutic drug targets. Scientific reports, 13(1):8366, May 2023.

[27] Ada Hamosh, Alan F Scott, Joanna S Amberger, Carol A Bocchini, and Victor A McKusick. Online mendelian inheritance in man (omim), a knowledgebase of human genes and genetic disorders. Nucleic acids research, 33(Database issue):D514—7, January 2005.

[28] Melissa J Landrum, Jennifer M Lee, George R Riley, Wonhee Jang, Wendy S Rubinstein, Deanna M Church, and Donna R Maglott. Clinvar: public archive of relationships among sequence variation and human phenotype. Nucleic acids research, 42(Database issue):D980—5, January 2014.

[29] FDA. Step 3: Clinical research — fda.gov. https://www.fda.gov/patients/drug-development-process/step-3-clinical-research, Apr 2018.

[30] Chi Heem Wong, Kien Wei Siah, and Andrew W Lo. Estimation of clinical trial success rates and related parameters. *Biostatistics (Oxford*, England), 20(2):273—286, April 2019.

[31] GitHub - gecko984/supervenn: supervenn: precise and easy-to-read multiple sets visualization in Python — github.com. https://github.com/gecko984/supervenn. [Accessed 07-06-2024].

[32] Open targets platform 23.12 has been released! https://blog.opentargets.org/open-targets-platform-23-12-release/#target-prioritisation, 2023. [Accessed 11-04-2024].

[33] Christian Fougner, Julie Cannon, Lydia The, Jeffrey F Smith, and Olivier Leclerc. Herding in the drug development pipeline. Nature reviews. Drug discovery, 22(8):617—618, August 2023.

[34] Duncan McElfresh, Sujay Khandagale, Jonathan Valverde, Vishak Prasad C, Benjamin Feuer, Chinmay Hegde, Ganesh Ramakrishnan, Micah Goldblum, and Colin White. When do neural nets outperform boosted trees on tabular data?, 2023.

[35] Yury Gorishniy, Ivan Rubachev, Valentin Khrulkov, and Artem Babenko. Revisiting deep learning models for tabular data, 2023.

[36] Gowthami Somepalli, Micah Goldblum, Avi Schwarzschild, C. Bayan Bruss, and Tom Goldstein. Saint: Improved neural networks for tabular data via row attention and contrastive pre-training, 2021.

[37] Roman Levin, Valeriia Cherepanova, Avi Schwarzschild, Arpit Bansal, C. Bayan Bruss, Tom Goldstein, Andrew Gordon Wilson, and Micah Goldblum. Transfer learning with deep tabular models, 2023.

[38] Noah Hollmann, Samuel Müller, Katharina Eggensperger, and Frank Hutter. Tabpfn: A transformer that solves small tabular classification problems in a second, 2023.

[39] Target - disease associations | Open Targets Platform Documentation. https://platform-docs.opentargets.org/associations#data-source-weights, 2023. [Accessed 11-04-2024].

[40] Charles Tapley Hoyt, Max Berrendorf, Mikhail Galkin, Volker Tresp, and Benjamin M. Gyori. A unified framework for rank-based evaluation metrics for link prediction in knowledge graphs, 2022.

[41] Alistair Moffat. Batch evaluation metrics in information retrieval: Measures, scales, and meaning, 2022.

[42] D Katz, J Baptista, S P Azen, and M C Pike. Obtaining confidence intervals for the risk ratio in cohort studies. Biometrics, 34(3):469, September 1978.

[43] Timothe Cezard, Fiona Cunningham, Sarah E Hunt, Baron Koylass, Nitin Kumar, Gary Saunders, April Shen, Andres F Silva, Kirill Tsukanov, Sundararaman Venkataraman, Paul Flicek, Helen Parkinson, and Thomas M Keane. The european variation archive: a fair resource of genomic variation for all species. Nucleic acids research, 50(D1):D1216—D1220, January 2022.

[44] Eleonora Porcu, Marie C Sadler, Kaido Lepik, Chiara Auwerx, Andrew R Wood, Antoine Weihs, Maroun S Bou Sleiman, Diogo M Ribeiro, Stefania Bandinelli, Toshiko Tanaka, Matthias Nauck, Uwe Völker, Olivier Delaneau, Andres Metspalu, Alexander Teumer, Timothy Frayling, Federico A Santoni, Alexandre Reymond, and Zoltán Kutalik. Differentially expressed genes reflect disease-induced rather than disease-causing changes in the transcriptome. Nature communications, 12(1):5647, September 2021.

[45] Chris Finan, Anna Gaulton, Felix A Kruger, R Thomas Lumbers, Tina Shah, Jorgen Engmann, Luana Galver, Ryan Kelley, Anneli Karlsson, Rita Santos, John P Overington, Aroon D Hingorani, and Juan P Casas. The druggable genome and support for target identification and validation in drug development. Science translational medicine, 9(383):eaag1166, March 2017.

[46] Chaohui Bao, Hengru Wang, and Hai Fang. Genomic evidence supports the recognition of endometriosis as an inflammatory systemic disease and reveals disease-specific therapeutic potentials of targeting neutrophil degranulation. Frontiers in immunology, 13:758440, 2022.

[47] Open Targets Tractability Pipeline (version 2). https://github.com/chembl/tractability_pipeline_v2, Jul 2023.

[48] Evan A Boyle, Yang I Li, and Jonathan K Pritchard. An expanded view of complex traits: From polygenic to omnigenic. Cell, 169(7):1177—1186, June 2017.

[49] Stephen Burgess, Amy M Mason, Andrew J Grant, Eric A W Slob, Apostolos Gkatzionis, Verena Zuber, Ashish Patel, Haodong Tian, Cunhao Liu, William G Haynes, G Kees Hovingh, Lotte Bjerre Knudsen, John C Whittaker, and Dipender Gill. Using genetic association data to guide drug discovery and development: Review of methods and applications. American journal of human genetics, 110(2):195—214, February 2023.

[50] Damian Szklarczyk, Rebecca Kirsch, Mikaela Koutrouli, Katerina Nastou, Farrokh Mehryary, Radja Hachilif, Annika L Gable, Tao Fang, Nadezhda T Doncheva, Sampo Pyysalo, Peer Bork, Lars J Jensen, and Christian von Mering. The string database in 2023: protein-protein association networks and functional enrichment analyses for any sequenced genome of interest. Nucleic acids research, 51(D1):D638—D646, January 2023.

[51] Robert W Hoffman, Joan T Merrill, Marta M E Alarcón-Riquelme, Michelle Petri, Ernst R Dow, Eric Nantz, Laura K Nisenbaum, Krista M Schroeder, Wendy J Komocsar, Narayanan B Perumal, Matthew D Linnik, David C Airey, Yushi Liu, Guilherme V Rocha, and Richard E Higgs. Gene expression and pharmacodynamic changes in 1,760 systemic lupus erythematosus patients from two phase iii trials of baff blockade with tabalumab. *Arthritis and rheumatology (Hoboken*, N.J.), 69(3):643—654, March 2017.

[52] Winston A Haynes, Aurelie Tomczak, and Purvesh Khatri. Gene annotation bias impedes biomedical research. Scientific reports, 8(1):1362, January 2018.

[53] Rita Santos, Oleg Ursu, Anna Gaulton, A Patrícia Bento, Ramesh S Donadi, Cristian G Bologa, Anneli Karlsson, Bissan Al-Lazikani, Anne Hersey, Tudor I Oprea, and John P Overington. A comprehensive map of molecular drug targets. Nature reviews. Drug discovery, 16(1):19—34, January 2017.

[54] Chris Finan, Anna Gaulton, Felix Kruger, Tom Lumbers, Tina Shah, Jorgen Engmann, Luana Galver, Ryan Kelly, Anneli Karlsson, Rita Santos, John Overington, Aroon Hingorani, and Juan Pablo Casas. The druggable genome and support for target identification and validation in drug development, 2016.

[55] Arielle Marks-Anglin and Yong Chen. A historical review of publication bias. Research synthesis methods, 11(6):725—742, November 2020.

[56] Luca Abatangelo, Rosalia Maglietta, Angela Distaso, Annarita D’Addabbo, Teresa Maria Creanza, Sayan Mukherjee, and Nicola Ancona. Comparative study of gene set enrichment methods. BMC bioinformatics, 10:275, September 2009.

[57] Katerina Trajanoska, Claude Bhérer, Daniel Taliun, Sirui Zhou, J Brent Richards, and Vincent Mooser. From target discovery to clinical drug development with human genetics. Nature, 620(7975):737—745, August 2023.

[58] Ruth J F Loos. 15 years of genome-wide association studies and no signs of slowing down. Nature communications, 11(1):5900, November 2020.

[59] Abdel Abdellaoui, Loic Yengo, Karin J H Verweij, and Peter M Visscher. 15 years of gwas discovery: Realizing the promise. American journal of human genetics, 110(2):179—194, February 2023.

[60] Michael J Bamshad, Deborah A Nickerson, and Jessica X Chong. Mendelian gene discovery: Fast and furious with no end in sight. American journal of human genetics, 105(3):448—455, September 2019.

[61] Olivier J Wouters, Martin McKee, and Jeroen Luyten. Estimated research and development investment needed to bring a new medicine to market, 2009-2018. JAMA, 323(9):844—853, March 2020.

[62] Clinical Development Success Rates and Contributing Factors 2011-2020 | BIO — bio.org. https://www.bio.org/clinical-development-success-rates-and-contributing-factors-2011-2020. [Accessed 31-07-2024].

[63] Joshua L Schoenbachler and Jacob J Hughey. pmparser and pmdb: resources for large-scale, open studies of the biomedical literature. PeerJ, 9:e11071, 2021.

[64] Direct and indirect evidence in Open Targets to expand associations. https://blog.opentargets.org/direct-versus-indirect-evidence-should-you-care/, 2017. [Accessed 11-04-2024].

[65] Konrad J Karczewski, Laurent C Francioli, Grace Tiao, Beryl B Cummings, Jessica Alföldi, Qingbo Wang, Ryan L Collins, Kristen M Laricchia, Andrea Ganna, Daniel P Birnbaum, Laura D Gauthier, Harrison Brand, Matthew Solomonson, Nicholas A Watts, Daniel Rhodes, Moriel Singer-Berk, Eleina M England, Eleanor G Seaby, Jack A Kosmicki, Raymond K Walters, Katherine Tashman, Yossi Farjoun, Eric Banks, Timothy Poterba, Arcturus Wang, Cotton Seed, Nicola Whiffin, Jessica X Chong, Kaitlin E Samocha, Emma Pierce-Hoffman, Zachary Zappala, Anne H O’Donnell-Luria, Eric Vallabh Minikel, Ben Weisburd, Monkol Lek, James S Ware, Christopher Vittal, Irina M Armean, Louis Bergelson, Kristian Cibulskis, Kristen M Connolly, Miguel Covarrubias, Stacey Donnelly, Steven Ferriera, Stacey Gabriel, Jeff Gentry, Namrata Gupta, Thibault Jeandet, Diane Kaplan, Christopher Llanwarne, Ruchi Munshi, Sam Novod, Nikelle Petrillo, David Roazen, Valentin Ruano-Rubio, Andrea Saltzman, Molly Schleicher, Jose Soto, Kathleen Tibbetts, Charlotte Tolonen, Gordon Wade, Michael E Talkowski, Genome Aggregation Database Consortium, Benjamin M Neale, Mark J Daly, and Daniel G MacArthur. The mutational constraint spectrum quantified from variation in 141,456 humans. Nature, 581(7809):434—443, May 2020.

[66] Mathias Uhlén, Linn Fagerberg, Björn M Hallström, Cecilia Lindskog, Per Oksvold, Adil Mardinoglu, Åsa Sivertsson, Caroline Kampf, Evelina Sjöstedt, Anna Asplund, IngMarie Olsson, Karolina Edlund, Emma Lundberg, Sanjay Navani, Cristina Al-Khalili Szigyarto, Jacob Odeberg, Dijana Djureinovic, Jenny Ottosson Takanen, Sophia Hober, Tove Alm, Per-Henrik Edqvist, Holger Berling, Hanna Tegel, Jan Mulder, Johan Rockberg, Peter Nilsson, Jochen M Schwenk, Marica Hamsten, Kalle von Feilitzen, Mattias Forsberg, Lukas Persson, Fredric Johansson, Martin Zwahlen, Gunnar von Heijne, Jens Nielsen, and Fredrik Pontén. Proteomics. tissue-based map of the human proteome. Science (New York, N.Y.), 347(6220):1260419, January 2015.

[67] Judith A Blake, Richard Baldarelli, James A Kadin, Joel E Richardson, Cynthia L Smith, Carol J Bult, and Mouse Genome Database Group. Mouse genome database (mgd): Knowledgebase for mouse-human comparative biology. Nucleic acids research, 49(D1):D981—D987, January 2021.

[68] Guolin Ke, Qi Meng, Thomas Finley, Taifeng Wang, Wei Chen, Weidong Ma, Qiwei Ye, and Tie-Yan Liu. Lightgbm: A highly efficient gradient boosting decision tree. In I. Guyon, U. Von Luxburg, S. Bengio, H. Wallach, R. Fergus, S. Vishwanathan, and R. Garnett, editors, Advances in Neural Information Processing Systems, volume 30. Curran Associates, Inc., 2017.

[69] James Malone, Ele Holloway, Tomasz Adamusiak, Misha Kapushesky, Jie Zheng, Nikolay Kolesnikov, Anna Zhukova, Alvis Brazma, and Helen Parkinson. Modeling sample variables with an experimental factor ontology. *Bioinformatics (Oxford*, England), 26(8):1112—1118, April 2010.

[70] Target - disease associations | Open Targets Platform Documentation. https://platform-docs.opentargets.org/associations#overall, 2024. [Accessed 11-04-2024].

